# Personality Change After Traumatic Brain Injury: A Systematic Review and Meta-Analysis

**DOI:** 10.64898/2026.06.29.26356816

**Authors:** Lauren Burns, Kelly Jones, Keishema Kerr, Nora Brennan, Natalie Clapshaw, Huw Green, Hannah Farrimond, Claire Stone, Sam Wilkinson, Members of Headway London, Vaughan Bell

## Abstract

**Background:** Personality change is a debilitating consequence of traumatic brain injury (TBI), yet its prevalence, characteristics, and treatment remain poorly understood.

**Methods:** We completed a pre-registered (CRD42023440990) systematic review and meta-analysis searching four databases (MEDLINE, PsycINFO, EMBASE and CINAHL) for primary studies assessing personality change after TBI. We synthesised conceptualisation, prevalence, longitudinal outcome, lesion location and treatment. Prevalence was estimated using a random effect meta-analysis using the Paule–Mandel estimator, with subgroup, meta-regression and robustness analyses.

**Results:** 101 studies were included in this review, seventeen of which were suitable for meta-analysis. Personality change was defined inconsistently although common symptoms involved the emergence or increase of affective, behavioural, and social disturbances, including irritability, depression, emotional instability, anger outbursts, social withdrawal, anxiety, impulsivity, restlessness, aberrant motor behaviours, and aggression. The prevalence of secondary personality disorder was estimated as 29.1% (CIs 22.5% – 36.2%) and prevalence of broad personality change was 68.1% (CIs 53.4% – 81.2%). Robustness analyses showed that the estimate for broad personality change should be treated with caution as it was unstable when adjusted for risk of bias and potential publication bias. Follow-up studies, although of varying quality, consistently showed personality change remained stable over long follow-up periods. The relationship between personality change and specific lesion locations in TBI remains unclear, likely due to the poor methodological quality of studies examining this association. Perhaps most concerning, there is limited evidence and very few systematic studies addressing treatment.

**Conclusion:** Personality change is a common and persistent consequence of TBI. Varying definitions, and the lack of high-quality lesion mapping studies and systematic investigations into treatment highlights critical gaps in understanding and management.

## Introduction

Traumatic brain injury (TBI) is a complex neurological condition caused by external mechanical forces that cause temporary or permanent impairment of brain function with wide-ranging physical, cognitive, and psychosocial consequences [1]. Personality change is considered one of the most disabling but least understood effects of TBI, affecting social adjustment, family life, and impacting rehabilitation outcomes [2].

Studies vary in how they define personality change including constructs spanning apathy to disinhibition [3]. As a result, it is often unclear which constructs are integral to personality change itself, as opposed to being part of broader cognitive or behavioural shifts. This diversity of definitions has led to various ways of classifying personality change making it hard to report comparable prevalences and, therefore, estimate the risk of personality change after TBI [4]. Patients and families affected by brain injury cite information about outcomes as important during the early stages of injury [5] but to date, there has been no systematic synthesis of the long-term prognosis of post-TBI personality change. Similarly, treatment options for secondary personality change have not been based on a systematic analysis of the literature [3] and it is not clear which treatments are used in those affected by post-TBI personality change. Finally, acquired personality change is traditionally associated with damage to the prefrontal cortex [6]. However, studies on other forms of acquired brain injury, such as tumour resection [7] and acquired brain injury more broadly [8], indicate that secondary personality change is associated with a far wider range of lesions. Understanding the diversity of lesions associated with post-TBI personality change may be useful in characterising the clinical presentation and neurocognitive basis of changes in affected patients.

Consequently, we completed a systematic review and meta-analysis to i) understand the most common symptoms associated with post-TBI personality change; ii) provide a meta-analytic prevalence of personality change after TBI; iii) review long-term outcomes and treatment, and; iv) review lesion locations associated with TBI-related personality change.

## Methods

The present review was undertaken following discussions with individuals with lived experience of brain injury from Headway London, a charity who support survivors of brain injury, who identified this as an important unmet need. The results were discussed and interpreted in partnership with them. The study is reported in compliance with the Preferred Reporting Items for Systematic Reviews and Meta-Analyses (PRISMA) 2020 guidelines and we pre-registered on PROSPERO (CRD42023440990).

### Types of studies included

We included all peer-reviewed primary studies published in English between July 1980 and November 2023 with the review updated in June 2025. We included studies using randomised or non-randomised controlled trials, retrospective or prospective cohort studies, case-control studies (including nested case-control and family studies), qualitative studies, and case studies, and excluded reviews, comments, newsletters, narrative book chapters, conference abstracts, and congress papers.

### Eligibility criteria

#### Participants

Studies investigating personality change following traumatic brain injury were included if they were original human research published in English with adult participants, while excluding reviews, non-English publications, animal studies, exclusively paediatric research, and studies of other acquired brain injuries. We originally defined ‘adult’ as 18+ but changed the inclusion criteria to 16+ following initial screening where 16+ was commonly used in TBI studies to define adult participation. Data from adults with TBI had to be distinguishable within a study if mixed populations were reported in a single paper.

#### Exposures

TBI was defined as any intracranial injury caused by an external force. This includes closed or penetrating head injuries, with severity ranging from mild to severe. Studies were excluded when general term such as “head injury” was used and the nature of the head injury could not be determined, or when TBI could not be differentiated from other acquired brain injuries such as stroke. Given the ambiguity in the definition of “personality change”, any papers that refer to “personality change” explicitly were included as were specific personality types (e.g. “narcissism”). Articles that identified acquisition of “personality disorder” diagnosis following a TBI were included. Any methods for assessing such changes were included. Studies were excluded if they solely discussed impulsivity, identity, cognition, behaviour, or emotion without specific mention of personality change.

#### Comparators

Studies with and without comparison groups were included; no exclusion criteria applied.

#### Outcomes

We included papers that reported the following outcomes: i) signs and symptoms associated with personality change following TBI; ii) diagnostic or classification tools used to assess post-TBI personality change; iii) prevalence of secondary personality change or personality disorder after TBI; iv) treatments for secondary personality change and their outcomes; v) lesion location associated with post-TBI personality change.

### Information sources and search strategy

Four databases were searched electronically (MEDLINE, PsycINFO, EMBASE and CINAHL). We conducted forward and backward reference searching for additional papers. See the Supplementary Material for full search terms.

### Study selection process

Review software Rayyan [9] was used to support the study selection phase, including duplicate removal, title and abstract screen, and full text screen. Duplicates were removed by one reviewer (LB). Two reviewers independently screened the title and abstracts, as well as full text articles (LB; KJ). A third reviewer (VB) was consulted when a consensus could not be reached during screening and full-text reading.

### Data extraction process

A spreadsheet was developed by one reviewer (LB) for extracting data from the full-text articles. One reviewer (LB) extracted study characteristics and outcomes of all included studies into the spreadsheet for comparison, which was conducted by two reviewers (LB; KJ), with a third held for disagreements (VB).

### Risk of bias

The quality of included studies using the Joanna Briggs Institute (JBI) critical appraisal tools. Study objectives, design, sampling methods, participant recruitment, measures, statistical analysis, results and conclusions were assessed for appropriateness. Disagreements were resolved either by consensus, or with the involvement of the third reviewer (VB).

### Synthesis of results

Prevalence was estimated using a random effect meta-analysis using the Paule–Mandel estimator implemented with the *meta* package [10] in *R* version 4.5.2 [11], with subgroup, meta-regression and robustness analyses. To assess publication bias, we used a Doi plot and the LFK index [12] due to funnel plots and Egger’s test being inappropriate for meta-analyses of prevalence [13]. A narrative synthesis was conducted to summarise symptoms reported as personality change, diagnostic and classification tools, treatment and long-term outcomes.

## Results

101 articles were included in the systematic review. See Figure 1 for PRISMA 2020 flow diagram. Studies were carried out in the USA (k=42), followed by the UK (k=18), Canada (k=7), Australia (k=4), Brazil (k=4), Denmark (k=4), Finland (k=3), China (k=3), Spain (k=2), and Taiwan (k=2). Belgium, Chile, Croatia, India, Ireland, Japan, Korea, New Zealand, Nigeria, Poland, Saudi Arabia, and Switzerland produced one study each. Study designs included cross-sectional studies (k=34), case studies (k=27), cohort studies (k=24), retrospective observational studies (k=7), case-control studies (k=3), case series (k=4), qualitative studies (k=1), and non-randomized intervention studies (k=1).

**Figure 1.**
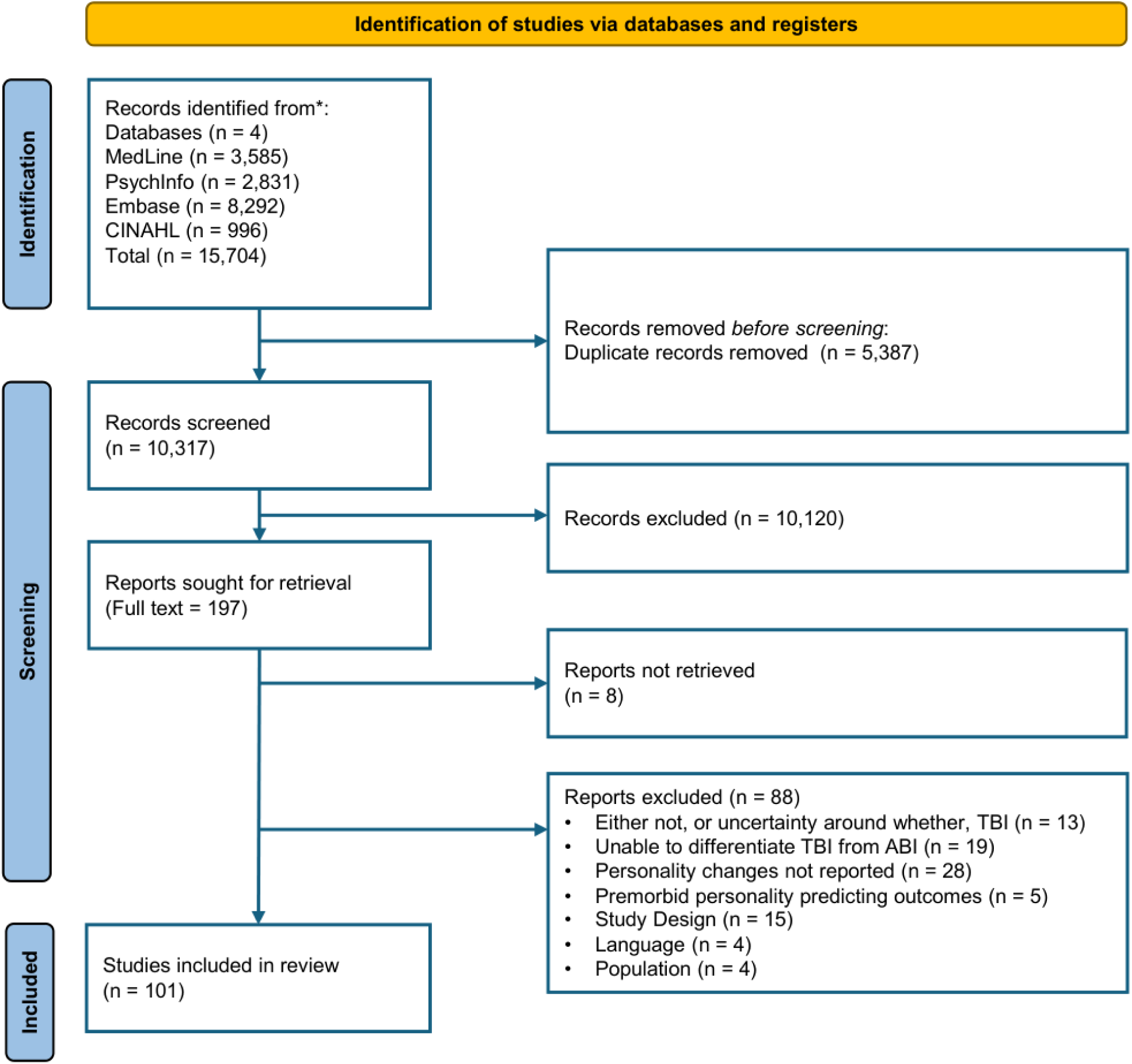
PRISMA 2020 flow diagram for systematic review

### Risk of bias assessment

We used the Joanna Briggs Institute (JBI) critical appraisal tool for risk of bias assessments. Details of the risk of bias ratings are reported in Figures S3a-S3d of the supplementary material. These indicated variable risk of bias, with higher risk of bias in cohort studies and inadequate control of confounding across studies designs.

### Synthesis of results

Table S2 in the supplementary material characterises all included studies. In additional to narrative synthesis, a meta-analysis was conducted with 17 studies reporting prevalence to estimate the prevalence of personality change following TBI. The analysis is available as a Jupyter Notebook, a document that combines code and the output in a form that can be re-run and reproduced, at the following URL: https://github.com/vaughanbell/personality-change-TBI-meta/

### Patient characteristics

The causes of TBI included road traffic accident (k=41), which included pedestrians, cyclists or vehicles. The second were falls (k=23), followed by assault (k=14), sports-related injuries (k=9), work-related injuries (k=9), gun-crime or war-related injuries (k=7). Nineteen studies reported “Other” categories, which included blows to the head, aircraft-related injuries, falling objects, or not otherwise specified. Thirty-eight of the studies did not specify causes of injury. The study populations were more frequently made up of men (male majority k=72, female majority k=13, unspecified k=12). Few studies reported ethnicity (k=15), although one study [14] found that ethnic minority individuals were disproportionally affected.

### Conceptualisation of post-TBI personality change

The signs and symptoms used in the conceptualisation of personality change by included studies are reported in Table 1, grouped under symptoms areas by the authors of this review. These have been included non-exclusively and signs and symptoms are reported as they were reported by the original authors. The most frequently reported signs and symptoms were irritability/frustration (k=21), depression (k=17), mood swings/emotional instability/emotionally impulsive (k=17), anger outbursts (k=14), decreased social interaction/increased social withdrawal/isolation (k=13), anxiety (k=12), impulsivity/impatience (k=11), restlessness/sleep disturbance/fatigue (k=10), aberrant motor behaviours/obscene gestures (k=10), and aggressiveness (k=9).

**Table 1.**
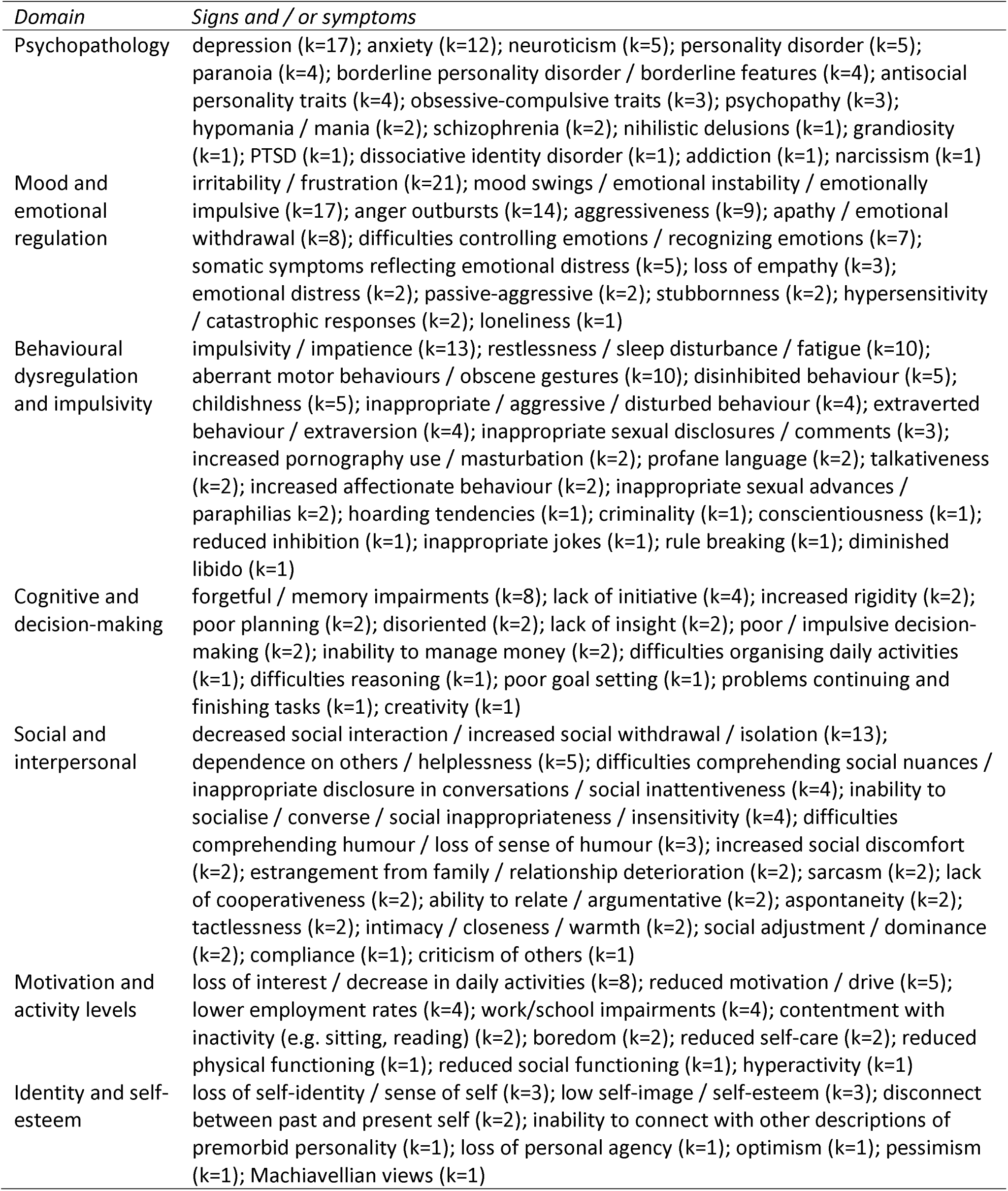
Signs and / or symptoms included under the conceptualisation of post-TBI personality change.

### Prevalence of post-TBI personality change

Among the 101 studies, 17 included prevalence data. An initial estimate of prevalence indicated an overall prevalence of 49.8% (CIs 37.3% – 62.3%) but with very high heterogeneity (I^2^ = 95.1%; tau^2^ = 0.0615l, p < 0.0001) indicating that this estimate is unlikely to be a valid synthesis of prevalence. A MetaForest analysis [15] to identify moderators that were the significant contributors to heterogeneity identified outcome type (broad personality change (k=9) vs diagnosis of secondary personality disorder (k=8) as the largest contributor to heterogeneity by a considerable margin (Figure S1, supplementary material) and subsequent analyses for broad personality change and diagnosis of secondary personality disorder were conducted separately. Figure 2 illustrates the forest plots for these meta-analyses.

**Figure 2.**
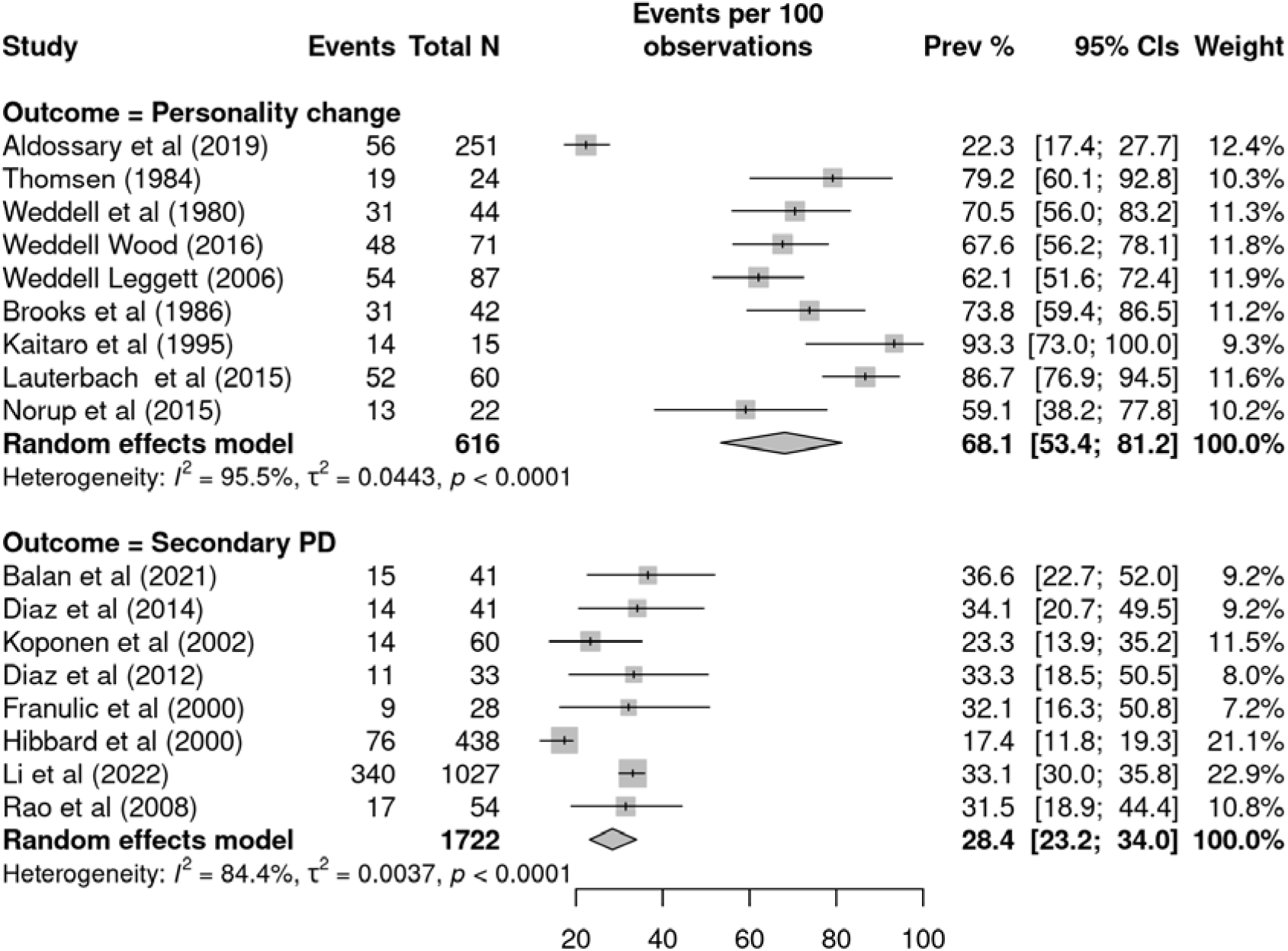
Forest plot of prevalence of post-TBI broad personality change and personality disorder diagnosis outcomes

#### Broad personality change

Broad personality change refers to personality change measured by studies using instruments or frameworks that extend beyond the specific changes captured within secondary personality disorder and included k=9 total studies. It was identified by relative report (k=3), clinical interview (k=2), SCAN (schedules for clinical assessment in neuropsychiatry; k=2), clinical review (k=1) and patient / relative questionnaire (k=1). Broad concept personality change prevalence was estimated as 68.1% (CIs 53.4% – 81.2%) with high heterogeneity (I² = 96%, tau² = 0.0443). However, outlier and leave one out sensitivity analyses identified Aldossary et al [16] as a significant outlier. Removal of this study led to a revised prevalence estimate of 73.7% (CIs 65.4% – 81.3%) with substantially reduced heterogeneity metrics (I^2^ = 65.4%; tau^2^ = 0.0095, p = 0.0101).

Doi plots and LFK indices were used to assess publication bias with evidence of asymmetry (LFK index = 2.26; Figure S2a, supplementary material) indicating the potential for publication bias affecting results. We used trim and fill to impute potentially missing small study effects, leading to an adjusted prevalence estimate of 42.5% (CIs 22.0% – 65.6%; I^2^ = 96.4%; tau^2^ = 0.1602). Subgroup analysis showed no statistically reliable moderation by longitudinal vs cross-sectional design or between studies that used validated measures vs unvalidated measures, although the latter comparison had only k=2 studies with validated measures. Meta-regression analyses showed a statistically reliable association with length of follow-up in months, although the effect was marginal increased prevalence (0.003; CIs 0.000 – 0.005) of personality change with each month of follow-up. There was no association between broad personality change prevalence and risk of bias assessment, although over the full sample there was an association with risk of bias lower prevalence was reported (−0.005; CIs −0.009 – 0.000; p = 0.037) with each percentage increase in risk of bias rating. Consequently, we calculated broad concept personality change prevalence adjusted by association with risk of bias rating across the whole sample, which gave a meta-regression adjusted prevalence of 51.2% (CIs 27.0% – 75.2%). We note the sensitivity of the broad personality prevalence estimate to adjustment, suggesting it is unlikely to be reliable and should be treated with caution. Full meta-regression results are reported in Table S3 of the supplementary material.

#### Secondary personality disorder diagnosis

The prevalence of secondary personality disorder was estimated as 28.4% (CIs 23.2% – 34.0%) with lower heterogeneity (I² = 84%, tau² = 0.0037). Outlier and leave one out sensitivity analysis identified Hibbard et al [17] as an outlier. Remove of this study led to a revised prevalence estimate of 32.5% (CIs 29.9% – 35.1%). Notably, the heterogeneity for this revised analysis was estimated as zero (I^2^ = 0%; tau^2^ = 0, p = 0.8214) although largely because removing Hibbard et al [17] left a single large study [18] and several small studies. The smaller studies contributed little weight to the overall between study variance, and all reported similar prevalence estimates, leading to these heterogeneity metrics.

Doi plot and LFK index (Figure S2b, supplementary material) showed no evidence of publication bias. Subgroup analysis showed no statistically reliable moderation by longitudinal vs cross-sectional design or between studies that used validated measures vs unvalidated measures. Meta-regression analysis showed an association with percentage of moderate TBI in the sample with more moderate TBI associated with lower prevalence (−0.007; CIs −0.013 – −0.001). There was no association between secondary personality disorder prevalence of risk of bias assessment. However, given the overall association in the full sample, we calculated the meta-regression adjusted prevalence of 29.5% (CIs 21.1% – 38.6%), differing only slightly from the original estimate. It was not possible to conduct a subgroup analysis on continent because study location was confounded with outcome type. Studies conducted in Europe were predominantly on broad concept personality change whereas those conducted in Latin and North American were predominantly on secondary personality disorder, with one study from Asia within each category.

### Longitudinal outcome

Longitudinal studies of personality change after TBI show little evidence for substantial change over time. However, studies were frequently small, with less than 50 participants. A prospective study by Diaz et al [19] followed-up 33 intensive-care-admitted brain injury patients 18 months after admission, reporting 33% of patients had secondary personality change. A small and retrospective study of 14 patients with complex maxillofacial trauma and high confidence of brain injury 2-11 years after injury by Snell et al [20] reported personality problems in 29-36% of patients. A study of 44 young adults with severe head injury conducted by Weddell et al [21] reported a 70% rate of clinician judged personality change at two years with higher rates of dependency, non-return-to-work, boredom and fewer interests in the personality changed group. Oddy et al [22] reported on the same cohort at 7-year follow-up and although there was no direct statistical comparison, the rate of personality change was almost unchanged. Two studies, Lezak [23] and Lezak and O’Brien [24], reported on the same cohort of 47 males with TBI over five years. Temperament and emotional issues remained stable after the first year, with 39–40% still affected at year five. Anger persisted, while anxiety and depression initially rose then declined, indicating that personality changes were not solely a reflection of comorbid affective disorder. Two studies relied on relatives’ ratings of the same group of severely head injured patients, reporting that perceived personality change increased from 60% at one year (N = 55) [25] to 74% at 5 years (N = 42) [26]. Kaitaro et al [27] directly compared relative and patient perceptions of personality change in an initial group of 19 severely head injured patients at 5 years, reporting that both groups perceived personality change to have occurred (patients 69%, relatives 80%) but there was almost no agreement on which personality traits had changed apart from agreement on a decrease in ‘excitability’. Thomsen [28] reported results from a long-term follow-up study of 40 patients with severe blunt head trauma, where prevalence had decreased from 80% at 2.5 years to 60% at 10-15 years. Only two papers reported longer follow-ups, albeit retrospectively and in single case studies. Both reported clear personality changes post-injury that persisted at 18 years [29] and 60 years [30] although with good social integration, largely due to support from the family and absence of sustained dissocial traits.

Studies using psychometric personality scales designed for the general population tended to report mixed results. A study of 54 patients with brain trauma reported that 31.48% scored in the abnormal range on the Korean Military Multiphasic Personality Inventory Test [31] although included patients with follow-ups ranging from 2 weeks to 12 years, making it difficult to judge what proportion were longer-term follow-ups. However, two studies using the Revised NEO Personality Inventory reported few changes at 3 and 12 months [32] and 30 days and 6 months [33].

A qualitative study of five women whose partners experienced personality change 1-7 years post-TBI reported [34] reported that even several years post-injury, three of five partners experienced their TBI spouses as fundamentally changed people, with relationship quality continuing to deteriorate over time rather than adapting positively.

### Lesion location

The evidence for reliable associations between lesion types and TBI-related personally change dimensions is poor, and there are few systematic studies. Table 2 reports lesions by personality change type reported in case studies. Frontal lobe involvement was common across personality dimensions. Temporal lobe lesions appeared more frequently in cases of irritability and aggression, while orbitofrontal involvement was most often reported with emotional lability and disinhibition. Bilateral lesions were relatively common in aggression and mood changes.

**Table 2.** Lesion location reported in TBI-related personality change case studies.

| *Personality dimension* | *N* | *Primary associated lesion location* |
| --- | --- | --- |
| Irritability / short temper | 11 | Frontal 73% (n=8), temporal 36% (n=4), right hemisphere 45% (n=5), bilateral 36% (n=4) |
| Mood changes | 10 | Frontal 60% (n=6), temporal 40% (n=4), parietal 20% (n=2), bilateral 40% (n=4) |
| Aggression / outbursts | 9 | Bilateral frontal 56% (n=5), temporal 44% (n=4), right hemisphere 44% (n=4) |
| Impulsivity | 5 | Frontal 100% (n=5), orbitofrontal/ dorsolateral prefrontal 60% (n=3), right hemisphere 40% (n=2) |
| Social disinhibition | 4 | Frontal 100% (n=4), orbitofrontal 50% (n=2), bilateral 50% (n=2) |
| Apathy | 5 | Frontal 100% (n=5), dorsolateral prefrontal 60% (n=3), right hemisphere 40% (n=2) |
| Emotional lability | 4 | Frontal 100% (n=4), orbitofrontal 75% (n=3), temporal 25% (n=1) |
| Social withdrawal / loss | 3 | Frontal 67% (n=2), temporal 33% (n=1), bilateral 33% (n=1) |
| Poor judgment inc. financial | 2 | Frontal 100% (n=2), bilateral 50% (n=1) |
| Anxiety-fearfulness | 2 | Frontal 50% (n=1), occipital 50% (n=1) |

However, substantial overlap, small sample sizes and potential reporting bias from single case do not permit firm conclusions regarding lesion–personality mapping.

Group studies used a range of methods for associating lesions with personality changes. Using the Eysenck Personality Questionnaire, Tate’s [35] study of 28 patients with varied lesion locations found consistent increases in neuroticism, addiction, and criminality scores regardless of specific damage sites. A study of 129 patients specifically associated these lesions with increased Machiavellianism tendencies with left dorsolateral pre-frontal cortex damage, suggesting this region may play a role in social-moral behaviour regulation [36]. A study of 10 patients with post-TBI organic personality disorder [37] associated predominantly frontal and temporal lesions with the ‘cognition energy’ and anxiety factors of the Neurobehavioural Rating Scale. Thomen’s [28] study included 40 patients, 15 with brainstem involvement, and 16 with focal lesions which were predominantly left sided (N = 12) with personality changes rated as childishness, emotional lability, irritability, restlessness and disturbed behaviour, but no specific analysis of lesion / personality change concordance.

### Treatment

#### Medication

We found no trials, controlled or uncontrolled, of medication to treat post-TBI personality change or secondary personality disorder. In a retrospective study, Llauterbach et al [38] reported medication used in the treatment of traumatic brain injury patients with severe neuropsychiatric symptoms including personality change. Out of 60 patients, N = 18 were prescribed mood stabilizers, N = 17 antipsychotics, N = 7 antidepressants, N = 3 benzodiazepines, N = 2 medroxyprogesterone, N = 2 beta blockers, and N = 2 medroxyprogesterone acetate to treat sexual aggression. Tsai and Hsiao [39] reported on a retrospective case series of 10 patients with secondary personality change in a cohort of psychiatric patients. They reported used of antipsychotics (N = 10) and carbamazepine (N = 7) to manage aggression, impulsivity, and expressiveness. In a single case study, carbamazepine was reported to ameliorate psychosexual behaviours in secondary personality disorder [40]. In another single case study, Margetic and Margetic [41] reported gradual improvements in personality traits (e.g., reduced anger and irritability) following quetiapine prescription.

#### Non-pharmacological interventions

Prigatano et al [42] reported that those attending a neurorehabilitation programme, involving intensive cognitive retraining and psychotherapeutic rehabilitation, compared to a control group, showed improvement in personality functioning compared to controls. In an uncontrolled study, Kaitaro et al [27] noted that rehabilitation was valued by families largely in terms of its benefits for changes in emotional reactions and personality. One case study [43] reported improvement in personality change after 20 sessions of repetitive transcranial magnetic stimulation, using 1Hz inhibitory stimulation on the right dorsolateral prefrontal cortex and 10Hz of excitatory stimulation on the left dorsolateral prefrontal cortex. We found no reports of the use of psychological interventions for post-TBI personality change.

## Discussion

This systematic review and meta-analysis found that personality change is common following TBI and tends to persist over long-term follow-up periods. However, literature shows significant variation in how personality change is defined, though certain common elements emerge across studies. The relationship between personality change and specific lesion locations remains unclear, likely due to the poor methodological quality of studies examining this association. Perhaps most concerning, there is limited evidence and very few systematic studies addressing treatment.

The meta-analytic synthesis of prevalence indicted that secondary personality disorder was present in 28.4% of patients post-TBI. When personality change was broadly defined, it was present in 68.1% of patients. However, the fact that this latter estimate changed markedly (42.5% when potential publication bias was adjusted for, 73.7% when an outlier was removed), with high heterogeneity in the original and adjusted figures, shows that this estimate is far less reliable and should be treated with caution. In contrast, the estimate prevalence estimate for secondary personality disorder remains broadly stable during robustness analysis, suggesting it is more likely to be reliable. Nevertheless, on conservative estimates, this suggests that roughly one third to one half of patients experience personality change post-TBI making it equally, if not more, common than cognitive impairment [44], and anxiety and depression [45] after TBI, although methodological heterogeneity across the studies included in these prevalence syntheses means that the prevalence estimates should be interpreted as broadly comparable rather than directly equivalent.

Post-TBI personality changes, as described in the literature, included a wide range of constructs. Some are commonly mentioned constructs in previous reviews of TBI-related personality changes [6,32] including changes such as irritability/frustration, anger, decreased social interaction, impulsivity and aggression. Nevertheless, depression, anxiety, poor sleep, and fatigue featured strongly in studies included in our review, and these are not typically considered personality characteristics either in the psychometric [46] nor clinical literature [47]. Our group of lived experience collaborators strongly advocated for fatigue and depression as being an important factor in personality change, due to its ability to limit social interaction and social vigour, and reduce tolerance for stress, leading to irritability. Psychometric studies of the general population [48] and studies in patients with stroke [49] report covariation between fatigue and personality. Personality appears to be relatively stable in patients with primary depression [50] although there is a larger degree of covariation between depression and personality disorder symptoms [51], indicating that depression may play a larger role moderating personality-related psychopathology, and may be an important factor in post-TBI personality change. These associations clearly need further investigation in post-TBI patients. We also note that personality change was described across the literature as including a surprisingly wide range of issues, including some psychiatric diagnoses and behavioural issues like addiction which are not typically considered to be in the domain of personality per se. Although these wider characteristics were typically mentioned in only a few studies, it may reflect the need to more effectively define post-TBI personality change to achieve a useful working consensus across clinical practice and research.

Longitudinal outcomes studies were typically small and of moderate quality but the results were broadly consistent across numerous studies, indicating that personality change remains prevalent after the post-acute period including in studies with long-term follow-ups of five [52], seven [22] or even ten or more years [28]. These findings suggest that personality changes following TBI are not only persistent but may represent a chronic condition that requires ongoing clinical attention and support. Nevertheless, the variability in measurement, sample size, follow-up periods, and inclusion criteria does suggest the need for better controlled studies to allow detection of trajectory-based subgroups, identify predictors of long-term outcomes, and clarify potential mechanisms.

Given this, we note the striking paucity of studies on interventions for TBI-related personality change. We found no studies which specifically targeted problematic personality change and few intervention studies that measured personality as an outcome. It is worth highlighting that individual treatment of problematic personality change is only one potential focus for treatment studies. Interventions for families to enhance support and adjustment are likely to be equally as valuable given evidence that TBI survivors and their family members may have poor agreement on how their personality has changed [27] and some partners report ongoing deterioration of their relationship in subsequent years [34]. The impact of change on family and relationship dynamics was highlighted by our lived experience collaborators as a priority for future research, as was the need for evidence-based therapies to support families affected by this.

Several factors might explain this gap. The wide range of characteristics included under the umbrella of TBI-related personality change, consensus definitions restricted to secondary personality disorder, and inconsistent measurement approaches, may discourage researchers from targeting it as a primary treatment outcome. There is a clinical tendency to treat component symptoms (such as irritability, impulsivity, or amotivation) rather than the overarching personality syndrome [53], which may perpetuate the absence of evidence for treating personality change as a coherent treatment objective. Qualitative studies suggest that personality change may be more likely to manifest as a problem after the affected person returns home,[54,55], meaning it may be missed by clinicians focusing on stabilisation. Additionally, outcome measurement remains a challenge. While a TBI-specific scale for personality change exists, [56] there is currently no research on which outcome measures are the most appropriate and most associated with longer-term functioning to guide intervention trials.

There was also little systematic evidence on lesion location and personality change. However, the results were broadly in line with a wider lesion mapping study that included patients with focal frontal lobe lesions not limited to TBI [8] where emotional lability was associated with orbitofrontal / ventromedial lesions and other frontal lesions were associated with disinhibited traits. Despite ‘disinhibition’ being widely used to describe personality changes, the extent to which they are best explained by frontal executive disinhibition remains poorly evidenced [57] indicating a need for future mechanistic research in this area.

We note some limitations of this review. Our inclusion criteria focused on studies that specifically addressed personality change, and some studies were excluded on the basis they addressed related concepts (e.g. disinhibition) but did not specifically address this in relation to personality as a broader concept. The extent to which these excluded constructs that overlap with, or meaningfully contribute to, broader conceptualisations of personality change, even if the authors of the original authors didn’t identify them as such, remains a matter of debate. In addition, diagnostic studies used a range of different measures, including structured assessments and clinical diagnoses, that may have relied on different diagnostic criteria as definitions are revised as new diagnostic systems emerge. It is unclear the extent to which this affects case identification and therefore prevalence estimates as it was not possible to test this statistically in this study. This study excluded studies not published in English, potentially limiting the inclusion of studies from countries and cultures where concepts and implications of post-TBI personality change may differ.

In conclusion, personality change is a common and enduring consequence of TBI, yet remains poorly addressed clinically. The development of targeted interventions for individuals and families affected by these changes remains a priority.

## Data Availability

All data produced are available online at:
https://github.com/vaughanbell/personality-change-TBI-meta/

## Author contributions

Conceptualisation: All authors; Methodology and Software: Lauren Burns and Vaughan Bell; Writing, original draft: Lauren Burns; Writing, reviewing: All authors; Screening: Lauren Burns, Kelly Jones and Vaughan Bell; Writing, reviewing: All authors; Writing, editing: Lauren Burns, Kelly Jones, Nora Brennan, Natalie Clapshaw, Huw Green, Hannah Farimond, Claire Stone, Sam Wilkinson and Vaughan Bell. Interpretation: All authors; Supervision: Vaughan Bell

## Availability of Data and Materials

All data and analysis code used in this study is available at https://github.com/vaughanbell/personality-change-TBI-meta/

