## Supplementary material for "Personality Change After Traumatic Brain Injury: A Systematic Review and Meta-Analysis"

**Figure S1.** Variable importance for MetaForest analysis identifying contributors to heterogeneity

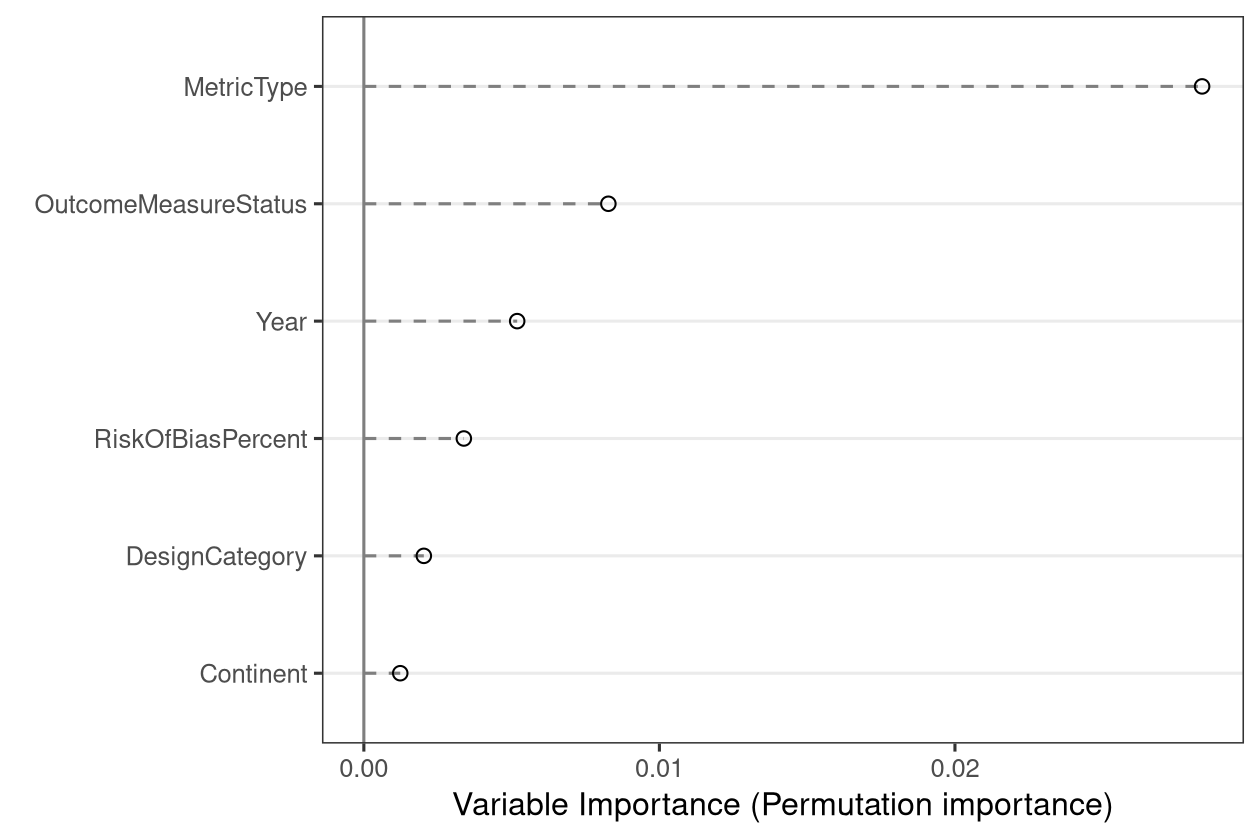

**Figure S2a.** Doi plot for broad personality change studies

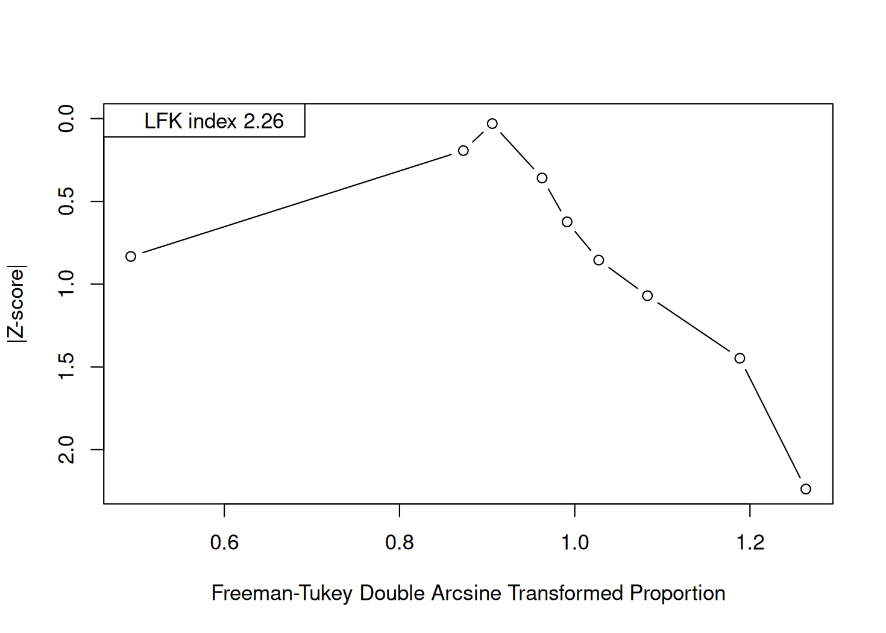

**Figure S2b.** Doi plot for secondary personality disorder studies

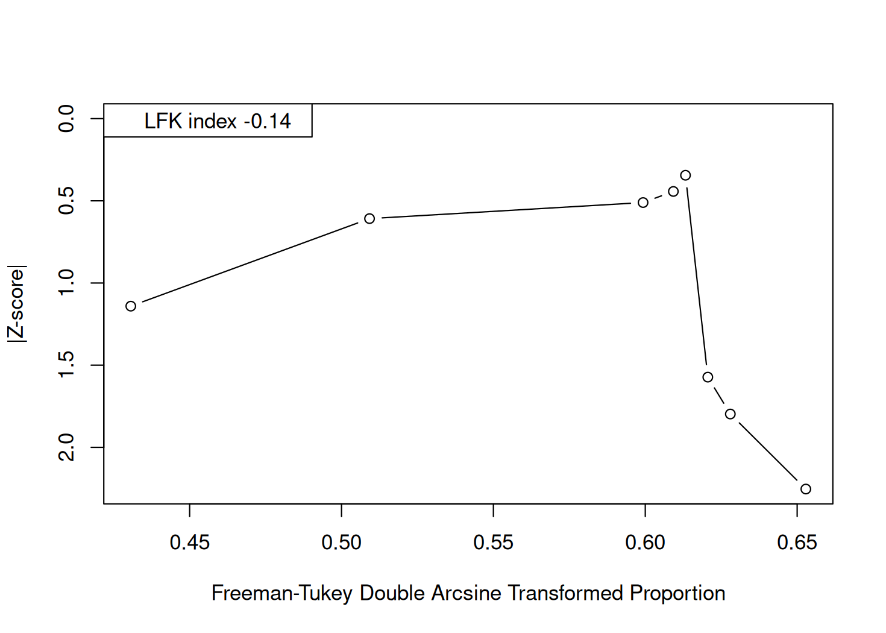

**Figures S3a.** Risk of bias ratings and risk of bias summary for cross-sectional studies

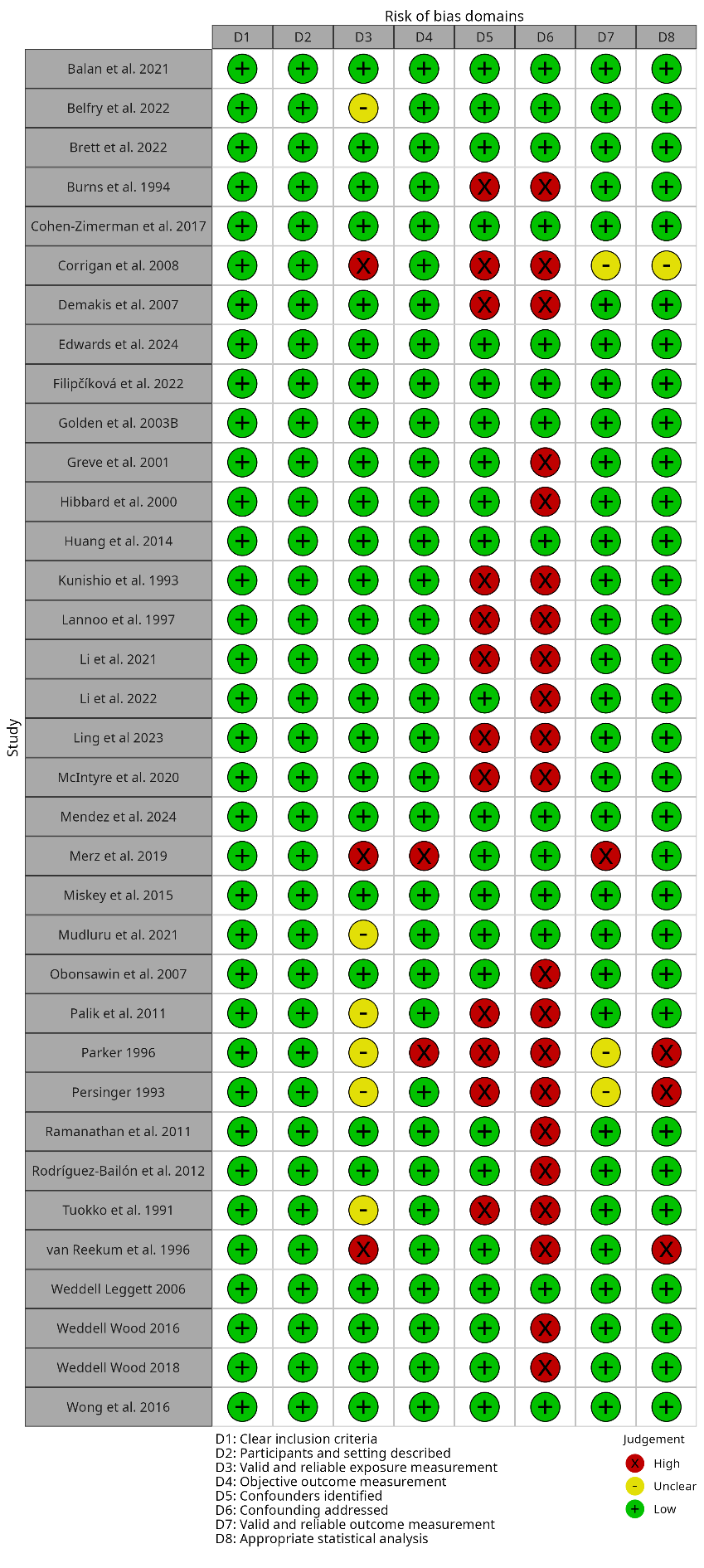

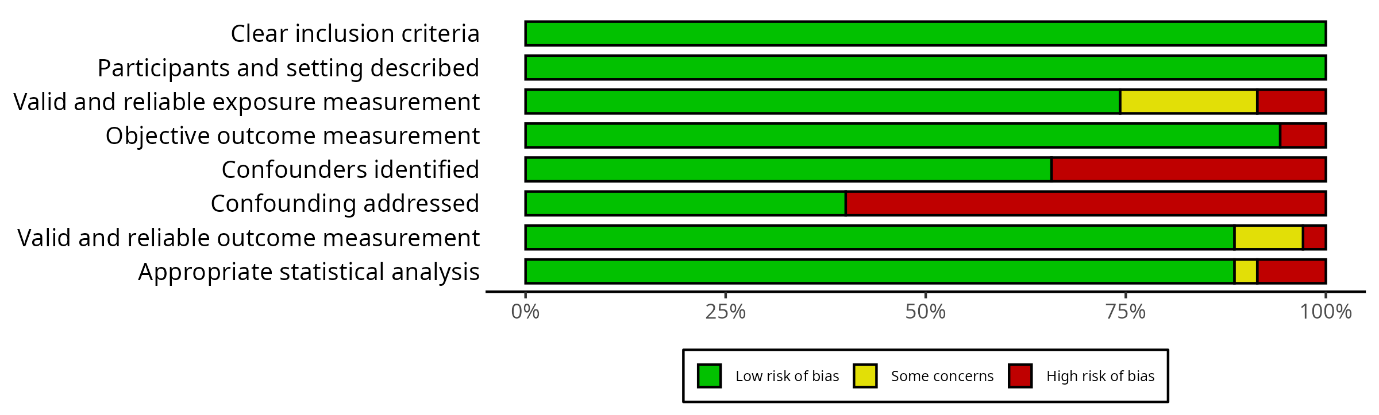

**Figures S3b.** Risk of bias ratings and risk of bias summary for cross-sectional studies

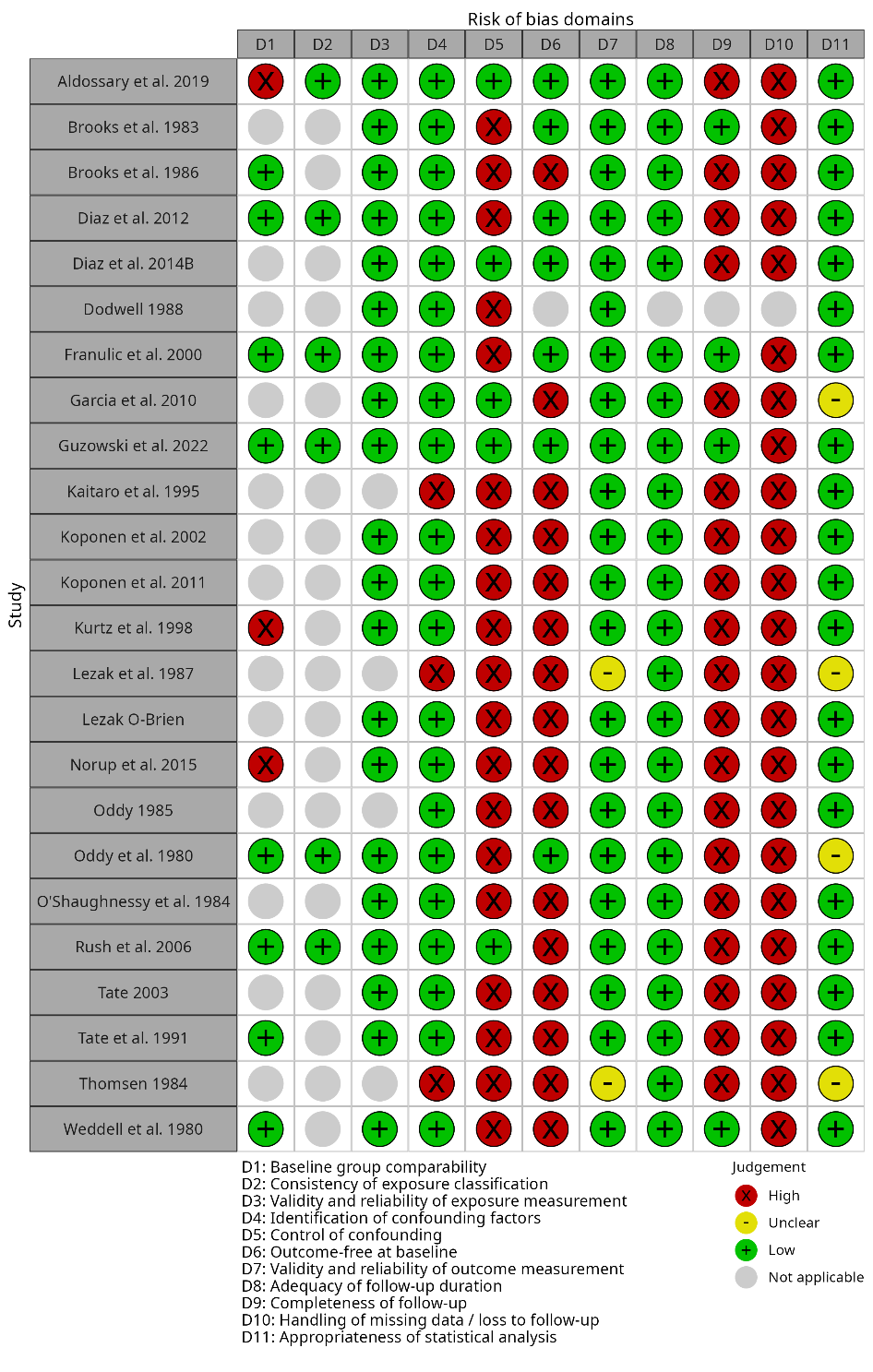

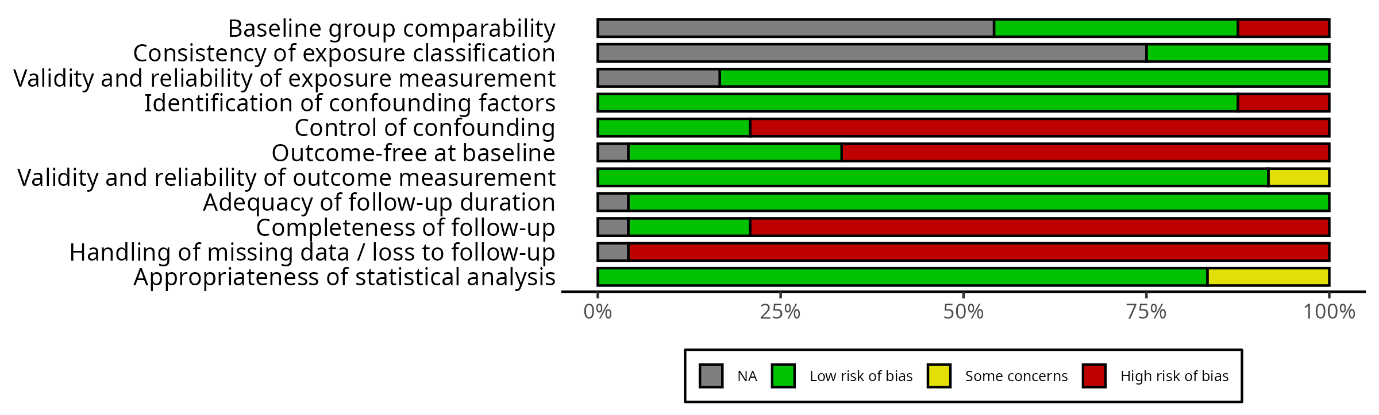

**Figure S3c.** Risk of bias ratings and risk of bias summary for retrospective observational studies

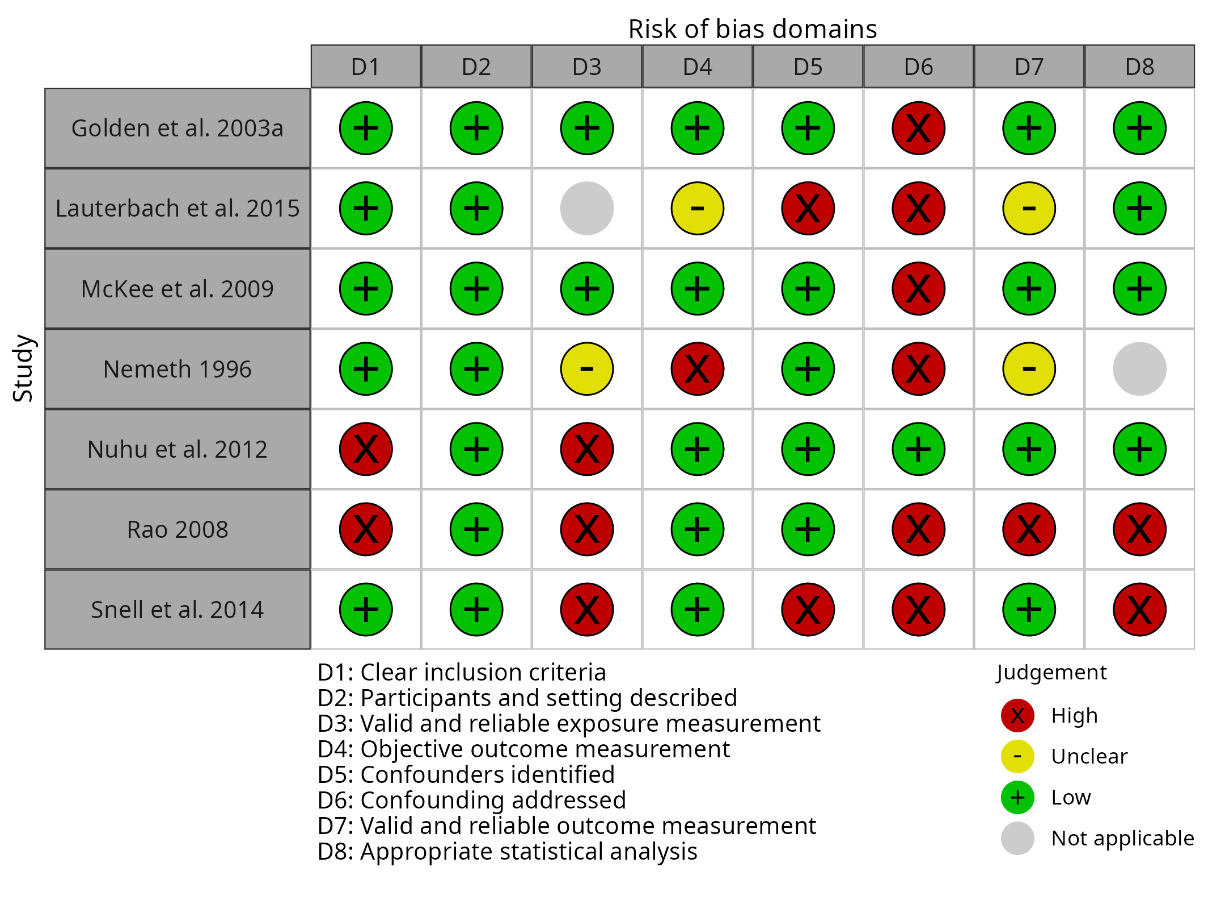

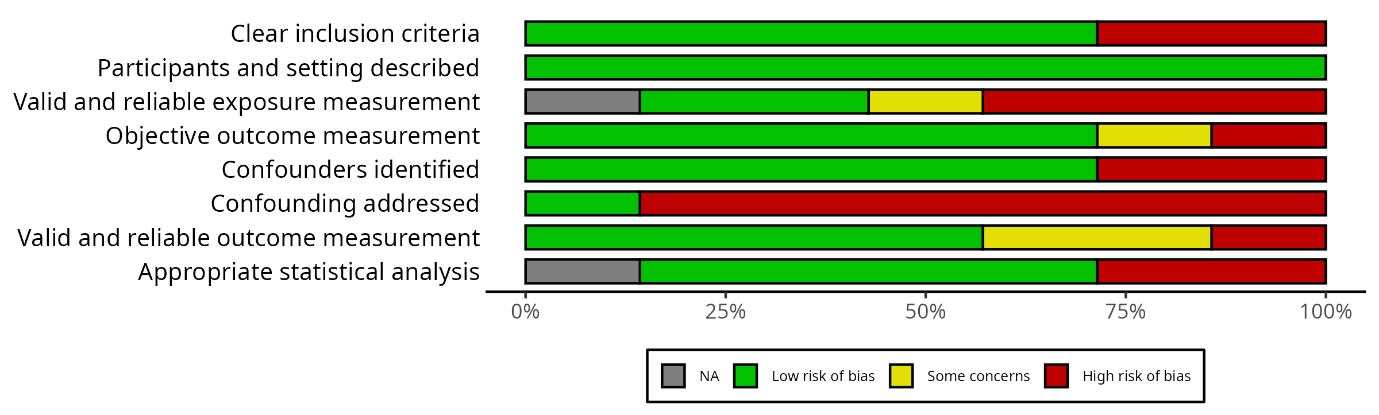

**Figure S3d.** Risk of bias ratings and risk of bias summary for case control studies

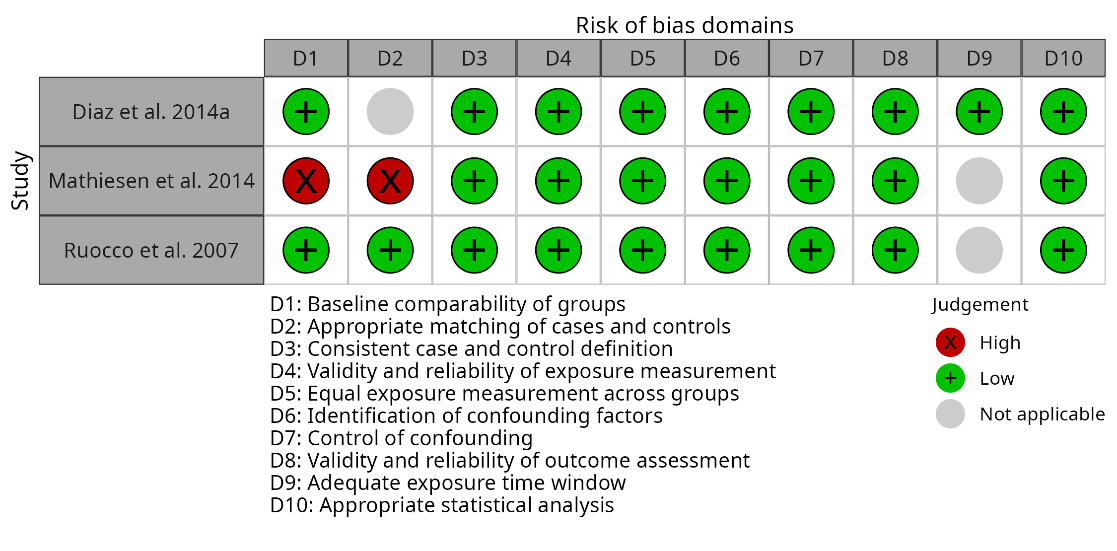

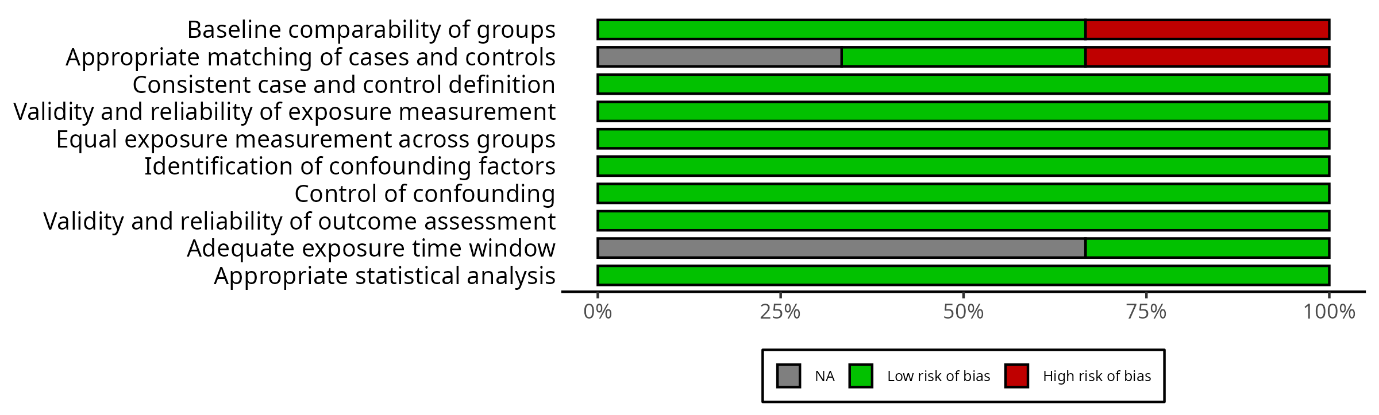

**Table S1**. Search strategy for the systematic review

S1 – Search Strategy

("traumatic brain injury" OR "TBI" or "craniocerebral trauma") AND ("personality" OR "personality change" OR "personality disorder" OR "personality syndrome" OR "frontal lobe syndrome" OR "prefrontal syndrome" OR "comportmental learning disabilities" OR "comportmental learning disability" OR "behavior disorder" OR "behaviour disorder" OR "behavioral disorder" OR "behavioural disorder" OR neurobehavi* OR antisocial OR dissocial* OR borderline OR histrionic OR narcissis* OR psychopath* OR sociopath* OR anankast* OR schizoid OR paranoid OR schizotyp* OR disinhibit*)

MeSH Terms: Brain Injuries OR Craniocerebral Trauma AND Personality

**Table S2**. Studies included in the systematic review

| **No.** | **Author** | **Year** | **Study Type** | **Participants** | **Country** | **TBI details** | **Personality outcomes** | **Instrument / Measures** | **Brain Injury Location** |
| --- | --- | --- | --- | --- | --- | --- | --- | --- | --- |
| 1 | Oddy & Humphrey | 1980 | Cohort study | TBIinitial=54, 6m=49, 12m=44, 2yr=39; CInitial=35, 6m=30, 12m=26, 2y=17 | UK | Closed head injury | Qualitative/Questionnaires | Kats Adjustment Scale | Not specified |
| 2 | Weddell et al. | 1980 | Cohort study | n=44, m=31, age=24.4, time between accident and admission = 6.4m | UK | Severe head injury | Post-injury: irritability {N = 17), increased affection (n=8), increased talkativeness (N = 4), increased disinhibition (TV = 3) and childishness (N = 4). | Oddy's interview; Bond Neurophysical Scale; Standard Progressive Matrices | Not specified |
| 3 | Thomsen | 1981 | Case study | Male, 44yo, edu=12y | Denmark | RTA | Partner gave pre-and post information | Neuropsych testing, but not specified | An X-ray revealed multiple fractures in the posterior left part of the skull |
| 4 | Goldberg & Buongiorno | 1982 | Case study | Male, 44 | USA | RTA | OPD | WAIS; Halsted Reitan battery | open skull fracture over the right frontal bone; prefrontal and frontal damage |
| 5 | Brooks & McKinlay | 1983 | Cohort study | TBI=55, M=46; age=m35.7 | UK | RTA=26; work-related injury=4; assault=11; other =14. | Personality adjective checklist: Present 3m=27, 6m=33, 12m=33; Absent 3m=28, 6m=22, 12m=22 | GCS | Not specified |
| 6 | O’Shaughnessy et al. | 1984 | Cohort study | N=39, M=29, age=28.2, edu=12.46, PTA=5.65days | USA | Closed head injury | EPI | WAIS; EMS; PASAT; TMT; EPI; Social Leisure Form | Not specified |
| 7 | Prigatano et al. | 1984 | Non-RCT intervention study | TBI=18, m=15, Age=26.1; C=17, m=15, age=23.5 | USA | Serious brain injury | KATS | WAIS; WMS; TMT; FTT; RN AIR; KATS | Severe Cerebral Contusion, Brain Stem Contusion |
| 8 | Thomsen | 1984 | Cohort study | n=40; m=28 age=21.5; f=12, age = 27.3 | Denmark | RTA=37; Falls =3; Closed head inj=9; Closed head inj + brainstem damage = 15; Closed head inj+verified focal lesion =16 | Changes in personality and emotion 2-5yrs=80%, 10-15yrs=65%: Childishness 2-15yrs=60% 10-15yrs=25%, Emotional lability = 40% 35%, Irritability 2-5yrs=38%, 10-15yrs=48%, Restlessness 2-5yrs=25%, 10-15yrs=38%, Disturbed behaviour 2-5yrs=23%, 10-15yrs=20%, Loss of social contact 2-5yrs=60%, 10-15yrs=68%, Aspontaneity 2-5yrs=43%, 10-15yrs=53%, Sensitivity distress 2-5yrs=23%, 10-15yrs=68%, Lack of interests 2-5yrs=20%, 10-15yrs=55% | WAIS | The worse overall outcome was seen in cases with severe brainstem involvement or anterior lesions or both. |
| 9 | Oddy et al. | 1985 | Cohort study | n=44, m=31 | UK | Closed head injury | Qualitative/Questionnaire | Bond Neuropsychological scale; ravens progressive matrices & mill hill vocab scale; Wimbledon self-report scale | Not specified |
| 10 | Brooks et al. | 1986 | Cohort study | TBI=55, M=46; age=m35.7. Relatives=42 | UK | RTA=26; work-related injury=4; assault=11; other =14. | Personality change 1year=60%, 5years=74% | GCS | Not specified |
| 11 | Nemeth | 1986 | Retrospective observational | n=21, m=10, 17to47age varying levels of education | USA | mTBI refers to a condition where cell loss or damage is assumed to have occurred as a result of a relatively mild cerebral concussion ; RTA=26, Falls=2, assaults=2, falling object=1 | Categorised as: 1. Impaired impulse control: Impatience, irritability, low frustration tolerance [n=18] 2. Inappropriate intensity of affect; mood swings [n=17] 3. Impaired motivation; loss of interest; social withdrawal; apathy. [n=17]. 4. Ability to relate [n=8]. 5. Diminished libido [n=6]. 6. Sleep disturbance [n=8] | GCS | Not specified |
| 12 | Lezak | 1987 | Cohort study | n=42, white, male, 27.8 | USA | RTA=28, Assault=7, falls=5, gunshot=1, airplane=1 | PAI | WAIS; PAI | Not specified |
| 13 | Dodwell | 1988 | Cohort study | n=56, M=46, age=26, O-levels=25, a-Levels=5, graduates=4 | UK | Blunt head injury | EPI | EPI; MNTS; MPI; PSE | Not specified |
| 14 | Lezak & O’Brien | 1988 | Cohort study | 42 white males with TBI (39 analyzed in some analyses). Mean age at injury: 27.1 years (SD=7.4), mean education: 12.4 years (SD=2.2). 17 unconscious <2 weeks, 25 unconscious >2 weeks. No prior psychiatric history. | USA | Unselected TBI sample. Etiology: 67% motor vehicle accidents, 17% blow to head, 12% fall, 5% other. Duration of unconsciousness: 8 patients <1 day, 9 patients <2 weeks, 23 patients >2 weeks, 2 patients >1 month. All examined within first year post-injury, followed for 5 years with 6 assessment periods. | 1) Persistent difficulties: anger (39% at year 5), anxiety, significant relationships (22-36%), social contacts, work/school, driving, appropriate social interaction; (2) Rapid improvement: initiative, residence, self-care, leisure, ambulation, aphasia; (3) Variable improvement: anxiety, depression. At 5 years, 45% displayed delusions/hallucinations or paranoia at some point. Emotional/social disturbances more handicapping than cognitive/physical disabilities. | Portland Adaptability Inventory (PAI) - developed for this study with 3 subscales: Temperament and Emotionality (7 items: anger, anxiety, indifference, depression, delusions/hallucinations, paranoia, initiative); Activities of Social Behavior (8 items analyzed: relationships, residence, social contacts, self-care, work/school, leisure, driving, appropriate social interaction); Physical Capabilities (6 items: ambulation, hands, auditory, vision, dysarthria, aphasia). 4-point scale (0-3). | Not specified |
| 15 | Tate et al. | 1991 | Cohort study | n=100; TBI=87, m=66, ageInj = 23.9, ageStudy=30, edu=9.3; C=40, m=28, ageStudy=29.2, edu=9.9 | Australia | Closed head injury = 58; Closed head injury with neurosurgery = 19; Open head injury = 20; RTA = 87% | In appendix A: PC TBI = 34 (40%), compliance =10, flexibility = 11, C+F =13; PC C = 3 (7.5%), compliance = 2, flexibility = 1 | Glasgow Outcome Scale | Not specified |
| 16 | Tuokko et al. | 1991 | Cross-sectional | N=79; age=33.9; time since inj=0.25 to 148 months mean = 16.27 months. | Canada | Not specified | MCMI | MCMI; GCS; WAIS | Not specified |
| 17 | Kunishio et al | 1993 | Cross-sectional | TBI=123, age=35.5, m=96; C=8 | USA | Focal or Diffuse injuries | Yatabe-Guilford personality test | WAIS; YGPT; GCS | Focal Injuries = acute epidural hematoma (EDH), acute subdural hematoma (SDH), in tracerebral hematoma (ICH), and low-density lesion.; Diffuse injuries= head injury accompanied by coma persisting for longer than 6 hours, but no CT evidence of a mass lesion, subdivided into diffuse axonal injury (DAI),') diffuse brain swelling (DBS), and hemispheric swelling (HS). |
| 18 | Persinger | 1993 | Cross-sectional | 56 neuro cases; TBI=39, m=23, C=17, m=10 | Canada | 90% RTA | Generalized activity, hedonism, arousal, and memory, were reported by all CHI patients but by none of the reference patients. | WAIS | frontal, temporal, temporofrontal and superior parietal lobes |
| 19 | Burns et al. | 1994 | Cross-sectional | n=85; m=71%, edu=13.1y, age at inj =34 y; age_test=36 y; time between inj and test = 2 y 9 m.; TBI=57, M=43, edu=13.; nTBI=28, M=17, edu=13. | USA | Traumatic brain injury (TBI) = 57 (inc assault i.e. gunshot wounds, blows; falls; RTAs) | Cattell's 16PF Personality test | LNNB; QAB; Cattell's 16PF Personality test | Not specified |
| 20 | Damasio et al. | 1994 | Case study | Phineas Gage - 25yo | USA | Work-related accident | Pre-injury: responsible, efficient, capable, intelligent, and socially well-adapted individual. Post injury: irreverent and capricious, lack of respect for social conventions, used profanities, lack of sense of responsibility. Unable to honour his commitments. | X-Ray | Left hemisphere: the anterior orbital frontal cortex, the polar and anterior mesial frontal cortices, and the anterior-most sector of the anterior cingulate gyrus. Right hemisphere: the anterior and mesial orbital region, the mesial and polar frontal cortices, and the anterior segment of the anterior cingulate gyrus. |
| 21 | Kaitaro et al. | 1995 | Cohort study | n=19, male=15, age inj=28, PTA=175days | Finland | Not specified | Not specified | WAIS, WMS, BVRT, | Not specified |
| 22 | Marshall et al. | 1995 | Case study | Male, 69yo | UK | RTA; work-related accident | his wife described her husband as a very gentle, calm man before the accident | MMSE | CT-scan showed bilateral frontal haemorrhages with additional right frontal lobe contusion. |
| 23 | Stratton & Gregory | 1995 | Case series | Case 1 = 31, ageinj=23, female; Case 2 = 18, ageinj=16, female; Case 3 = 31, ageinj=29, female; Case 4 = 19, female | New Zealand | RTA = 1, 2,3,4 (pedestrian/passenger) | Case 1 = no ref to personality change; Case 2 = no longer had any sense of humor and jokes went over her head. Pre-injury: good sense of humour.; Case 3 = Severe depression, loss of confidence, irritability and impatience. Pre-injury: optimistic person with a feeling of well-being and happiness.; Case 4 = Reported no personality changes. | Not specified | Case 1 = left frontal temporal and occipital areas; Case 2 = Frontal lobe; Case 3= Occipital areas; Case 4= posterior temporal region, right hemisphere |
| 24 | Parker | 1996 | Cross-sectional | Total = 33; edu = 14.2 years; time since inj=20 months, age=38.5 | USA | RTA | A wide spectrum of disorders was observed: cerebral personality disorder, persistent altered consciousness, post-traumatic stress, psychodynamic reactions to impairment, and complex reactions expressing neurological, somatic, and psychological dysfunctions (sexuality and somatization). | WAIS; Inkblots; Figure drawings | Not specified |
| 25 | Van Reekum et al. | 1996 | Cross-sectional | n=18; m=8; post-inj=4.9y; 19-49y | Canada. | sevTBI=10, modTBI=8; | DSM-IV | SIDP-R; SADS-L | Not specified |
| 26 | Harrington et al. | 1997 | Case study | Male, 59yo, Israeli born | USA | Fall | Post-injury: disinhibited behavior, mood swings, delusional jealousy, temper outbursts, impulsivity, foul language, decreased hygiene and self-care and decreased interest in his usual activities. He was easily fatigued and very impatient. | Not specified | bifrontal hemorrhagic contusions right > left, with subfalcine herniation from right to left and downward mass effect with blood in the right lateral ventricle. A right frontal subdural hematorna. Small, old lacunar infarction in the left caudate nucleus/ anterior limb of the internal capsule. |
| 27 | Labbate et al. | 1997 | Case series | A=Male, 20yo; B=Male, 34yo; C=Female, 29yo | USA | A= Work-related accident; B=RTA; C=RTA | A= Before was very anxious socially and urinating. Now, he longer felt uncomfortable with groups and felt more confident, able to speak his thoughts. He rarely blushed and was able to urinate without difficulty for the military mandatory urine drug tests. Seven months after the injury much of his irritability and emotional lability had resolved. He, however, continued to feel confident and noticed the absence of anxiety persisted in all social situations.; B= His former wife states that around September 1976 he had personality change. Prior to the injury, he had antisocial personality disorder. He no longer has sociopathv interests since the motorcycle trauma.; C= Following the injury she reported mild memory deficits, but noticed a marked change; she was no longer always angry. Two months after the injury, coworkers and friends in the Navy also noticed that she was much more calm and friendly, she was able calmly to complain about concerns with ward nurses, whereas formerly she commonly became extremely angry in such instances. She was not apathetic, and her mood was euthymic. | A=WAIS, WCST, DSM-V ("personality") | A=No abnormality; B= a right hemothorax, an acute subdural hematoma in the right cerebral hemisphere, and cerebral edema, and he underwent a bifrontal craniotomy. |
| 28 | Lannoo et al. | 1997 | Cross-sectional | TBI=68, m=54, age=35, edu=11; Trauma=28 m=20, age=37, edu=12 | Belgium | Moderate-Severe head injury | NEO-FFI | NEO-FFI; GCS | Not specified |
| 29 | Kurtz et al. | 1998 | Cohort study | TBI=21, m=18, black=13, age=32.71; C=25, m=18, black=11, age=35.48 | USA | RTA=12, assault=6, bicycle=2, fall=1 | NEO-PR-IV | NEO-PR-IV; REC | Not specified |
| 30 | Cantagallo et al. | 1999 | Case study | Male, 32yo | UK | RTA | Acquired DID post TBI | Not specified | CT Scan did not disclose any abnormality. EEG saw patchy dyshomogeneous uptake with left temporal and biparietal hypo- perfusion, worse in the left hemisphere. |
| 31 | Dimitrov et al. | 1999 | Case study | Male, 50yo (injury sustained at 20yo) | USA | War (Shrapnel) | Post-injury: Unusual moodiness, mood swings, sarcasm, lack of tact with others, bluntness of affect, remoteness of rapport, social withdrawal, no ability to make and keep friends, and, especially, questionable competency in handling large sums of money, unable to plan daily activities…. He “possessed the experience and ethics of a 14 year old”, increased pornography use. | WAIS, WMS, BADS, WCST, BDI | right frontal ventromedial lesion |
| 32 | Neylan | 1999 | Case study | Phineas Gage | USA | Work-related accident | Pre-injury: efficient, capable.; Post-injury: fitful, irreverent, profane, unempathetic, impatient, obstinate, capricious and vacillating. | Not specified | Right prefrontal cortex |
| 33 | Tsai et al. | 1999 | Case series | n=10, TBI=6/10, age=37.6 | Taiwan | Head trauma | Affective liability =5; aggression =4; impulsivity =2; grandiosity =1; impulsivity =3; paranoia=3; apathy=2 | DSM-IV | Not specified |
| 34 | Franulic et al. | 2000 | Cohort study | w/OPD n=9, m=0, age=41av; w/o OPD n=21, m=3, age=33av | Chile | TBI | ICD-10 defined organic personality disorder | ICD-10; WAIS, Benton TestROT, WCST, PS, NRS-27. | OPD = 6 x disturbed cerebral TC, specifically frontal and temporal secondary type. The most frequent lesion was of the haemorrhagic contusive type. |
| 35 | Glover | 2000 | Case study | Male, 34yo | UK | Fall | Pre-injury: vibrant, energetic physical man.; Post-injury: “a shell of himself”, angry, frustrated, short tempered, unable to reason, argumentative, stubborn. | Not specified | Not specified |
| 36 | Hibbard et al. | 2000 | Cross-sectional | n=100, TBIw/oPD=76, TBIw/PD=24, m=53, age=38, White=73, Black=14, Hispanic=9, Other=4 | USA | Motor vehicle accident 62%; Assault 8%; Pedestrian accident 8%; Fall 6%; Sport related 6%; Hit by falling or flying object 5%; Gunshot wound 2%; 41% Other means 3% | DSM-IV Criteria | DSM-5 ("personality change"); | Not specified |
| 37 | Greve et al. | 2001 | Cross-sectional | IA n=26, m=24, age=33.9, education=11.4, injuryage=23.3, comalength=44.85; C=19, m=17, age=38.9, education = 12.9, injuryage=26.8, comalength=42.9 | USA | TBI | Eysenck Personality Questionnaire (EPQ) | LHA, BPAQ (aggression); EPQ (personality); BIS (impulsivity); PPVT, TMT, COWAT, WCST | Not specified |
| 38 | Mataró et al. | 2001 | Case study | Male, 81 (inj=21) | Spain | Fell onto an iron stake | Post-injury: dependent on others, cheerful, problems establishing realistic goals, lack of drive, difficulties initiating, continuing and finishing activities. Lack of drive and concern, apathy, poor planning. | Not specified | Bilateral damage affecting the orbital and dorsolateral frontal regions |
| 40 | Koponen et al. | 2002 | Cohort study | n=60; male=41, age=60.8, ageinj=29.4, | Finland | Not specified | DSM-IV Criteria | DSM-5; MMSE | Not specified |
| 41 | Tate | 2003 | Cohort study | Relatives Initial = 45, 6m=30, 12m=28; TBI; m=24 (85.7%), age=26.82, edu = 10.29 | Australia | RTA =21 (75%), assault = 5, fall =1, sport (soccer) = 1. | EPQ-R | EPQ-R; CBS | structural lesions in frontal lobes = 17 |
| 42 | Golden & Golden | 2003.1 | Retrospective observational | 320 TBI pts; Ethnicity - Caucasian (82.2%), Black (14.1%), Hispanic (3.8%). M=234 (73.1 %), age 45.4 (SD = 12.69), education 12.02 (SD = 2.012) | USA | TBI | MMPI | MMPI-2; HRNB | Not specified |
| 43 | Golden & Golden | 2003.2 | Cross-sectional | Stroke n= 124. HI = 290. Dementia n= 66 ; Av 58.04yo, education = 12.67, mostly Caucasian but 80 African-Americans, Hispanics, or others. Av chronicity = 77.55 months | USA | TBI | MMPI | MMPI-2 | Not specified |
| 44 | Heinrich & Junig | 2004 | Case study | Male, 69yo | USA | RTA | Post-injury: irritable, slightly elevated. Noted changes. Thought process was tangential and difficult to redirect at times. | Folstein Mini-Mental Status Examination | left-sided frontal bacute on chronic SDH with mild left frontal mass effect. A left frontal SDH and gliosis in the left orbitofrontal and left posterior basotemporal lobes along with the right occipital lobe (contra-coup injury). |
| 45 | Wood & Rutterford | 2004 | Case study | Male, 40yo, age inj=22 | UK | RTA | Pre-injury: quiet, mild mannered person. Post-injury: disorientated, restless, and agitated. Post-operatively, more outspoken, less agitation, bright, cheerful and articulate. 9months: Irritability, agitation, poor concentration, difficulty with memory, inordinate fatigue, lethargy, and a poor sleep pattern. 15 months: extraverted, tiredness. lacked inhibition and social judgement, told socially inappropriate jokes or comments. | WCST | Bifrontal damage. Walls of the frontal sinuses were extensively fractured, with damage to the ethmoidal complex and right orbital wall. Breach in the dura near the midline on the left side. Cerebrospinal fluid rhinorrhoea was present. |
| 46 | Chang et al. | 2006 | Case study | Male, 77yo | Taiwan | Fall | Change in personality characterized by bad temper, disturbed sleep patterns, easily fatigue ability and poor appetite | Glasgow Coma Scale | mild subdural effusion in bilateral frontotemporal areas. Cerebrospinal fluid was normal. Biochemistry studies showed only mild hyponatremia and mild hypokalemia |
| 47 | O’Gorman | 2006 | Case study | Male | UK | RTA | Pre-injury: laid back, easy–going, creative.; Post-injury: Frightening dreams, angry outbursts, out of control emotionally. | Not specified | cerebral contusions – in both frontal lobes and brainstem areas of the brain |
| 48 | Rush et al. | 2006 | Cohort study | n=90, mTBI=20; Mod-SevTBI= 39; C=3. Ax Data= 38-56 days post-inj, 1-2yr follow up= 391-442 days post-inj | USA | GCS | NEO-PI-R | NEO-PI-R (personality), GCS; GOAT; MPAI; ILS; VIS. | Not specified |
| 49 | Weddell & Leggett | 2006 | Cross-sectional | n= 87; M=71; age=? | UK | Severe TBI=72; Mild TBI=15 | noPC (n = 24), PC? (n = 9) and yesPC (n = 54) | GCS; WAIS; WMS; UPSIT; STA; ZDS; GHQ-28 | orbitofrontal and/or medial temporal damage |
| 50 | Demakis et al. | 2007 | Cross-sectional | Combined= 95, age=33.4, edu=12.5, m=82.1%, w=72%; Rehab =60, age=36.8, edu=12.4y, m=73.3%, w=80%; Military= 35, age=27.5, edu=12.5, m=97.1%, w=57% | USA | Rehab: RTA=73%, falls=15%, other=12%. Military: blast=57%, other =43% | PAI | PAI | Not specified |
| 51 | Handel et al. | 2007 | Case study | Male, 58yo | USA | RTA | Post-injury: Withdrawn, excessively worried about money, nihilistic delusions about his lack of resources and convictions about impoverishment. Decline in mood, sleep, appetite, interest in self-care, and participation in daily activities. Stopped working. | WAIS | subarachnoid hemorrhage, primarily involving the left fronto-parietal regions |
| 52 | Obonsawin et al. | 2007 | Cross-sectional | TBI=184, m=152, age=35.2, age inj=28.8; C=87, m=57, age=35.8 | UK | Not specified | 123 personality characteristics gathered from (1) literature on TBI, (2) personality assessment scales, (3) rehabilitation staff (4) clients (5) the Scale for the Assessment of Negative Symptoms14 and the Scale for the Assessment of Positive Symptoms,15 and (6) literature on personality disorder. | GCS; SANS; SPNS | Not specified |
| 53 | Ruocco et al. | 2007 | Case control | Total=462; TBI=231, age=40.6, edu=13.6yrs, m=128; Psychiatric=231, age=40.9, m=128, W=87, AA=29, H=11, A=3, edu=13.6 | USA | RTA =74.8%, falls =11%, blows to the head =10.6%, physical assault =2.2%, aircraft crash =1.3%. mildTBI=84%, moderateTBI=16% | MCMI | MCMI | Not specified |
| 54 | Corrigan & Deutschle | 2008 | Cross-sectional | n=50; m=31, w=92%, age=37.5; less than high school diploma=44%, 42% = diploma or GED; 14% = college or advanced training. | USA | COFD item = ‘Have you ever hurt your head or had a head injury that resulted in being knocked out or being taken to the hospital?’ | Axis II Personality disorder diagnoses. Borderline personality disorder TBI=4 (11%), nonTBI=0 (0%); Antisocial personality disorder TBI=2 (6%), nonTBI=0 (0%); Personality disorder. NOS TBI=3 (8%), nonTBI=0 (0%); Any personality disorder TBI=9 (25%), nonTBI=0 (0%) | COFD |  |
| 55 | Namiki et al. | 2008 | Case study | Male, 54yo (inj @53) | Japan | RTA | Post-injury: disinhibition relating to impulsivity and sexual matters. Easily angered or irritated, emotional outbursts, makes inappropriate sexual comments and advances, is too flirtatious, is hyperactive and unable to sit still. | FrSBS, WAIS, WMS, BFRT, WCST, BADS | Ambiguous findings in the orbitofrontal cortex area were found when using the MRI, EEG or SPECT with N-isopropyl-p-[123I] iodoamphetamine (IMP). |
| 56 | Rao et al. | 2008 | Retrospective observational | Total=54; age=41.1 years; male=37; w=81.5%; edu=13.4years. | USA | Closed Head-Injury; 44%=mTBI, 20.4%= moderate, 35.2% sTBI. | DSM-IV Criteria | GCS; MMSE | Not specified |
| 57 | Dasarathy et al. | 2009 | Case study | Female, 82yo | UK | RTA | Pre-injury: pleasant and polite. Post-injury: agitation, verbal aggression and expression of obscene gestures. | MMSE | Contra-coup bilateral frontal contusions and small sub-arachnoid, subdural and extradural haemorrhages |
| 58 | McKee et al. | 2009 | Retrospective observational | n=51, m=49, SymptomAge=42.8, age starting=15.4; Case A=45yo, white male; Case B = 80yo, african american man; Case C=73yo, male | USA | Sport-related injury; boxing=39, american football=5, wrestler=1, soccer=1. | Case A = NFL linebacker, became angry, verbally aggressive, more emotionally labile.; Case B = boxing for 5 yrs, wife referred to him as "punch-drunk" in mid-30s, and at age 78 became paranoid; Case C=boxing, late 50s became forgetful, mood swings, restless, apathetic, socially withdrawn, paranoid, irritable, and sometimes violently agitated. Previously happy, easy-going self. | Not specified | Case C: Computed tomographic scan and magnetic resonance imaging (MRI) showed generalized cortical atrophy, enlargement of the cerebral ventricles, cavumseptum pellucidum, and a right globus pallidus lacuna. |
| 59 | Gracia-Garcia et al. | 2010 | Cohort study | 41 pts | USA | TBI | personality profiles were captured with the 60-item NEO Five-Factor Inventory (NEO-FFI) | NEO-FFI | Not specified |
| 60 | Koponen et al. | 2011 | Cohort study | n=38, m=27 (71.1%), ageinj=41.6, edu=10.7 | Finland | Fall n=26 (68.4%); RTA=10 (26.3%); Assault =1 (2.6%); Other cause =1 (2.6%); Mild = 27 (71.1%); Moderate = 6 (15.8%); Severe = 3 (7.9%); Very severe = 2 (5.3%) | SCID | SCAN; SCID | Not specified |
| 61 | Paik et al. | 2011 | Cross-sectional | Total = 237; Contol = 150, BT=54, BD=33, male=100%, 19yo, brain trauma = 2 weeks to 12 years ago | Korea | Not specified | KMPI | KMPI | Not specified |
| 62 | Ramanathan et al. | 2011 | Cross-sectional | Total =45; M=25; W=40, B=4, A=1; age=34.1;edu=13; | USA | Not specified | Not specified | SCL-90-R; TICKS; CHARTS; LOT-R | Not specified |
| 63 | Colantonio & Comper | 2012 | Case series | Male = 75%, > 44 yo = 69%, married = 70%, less than high school education (47%). | Canada | Work-related accident | Not specified | RPQ | Not specified |
| 64 | Diaz et al. | 2012 | Cohort study | Total n =48; male = 42, mean age = 32.33.; No Psych Dis = 15; male = 13, mean age = 34.45; Psych Dis = 33; male = 29, mean age = 31.36 | Brazil | Severe TBI | Thus, for this study, ‘‘personality changes’’ is considered to be personality changes in general, more specifically due to a severe head trauma, reported by family. | Glasgow Coma Scale; DSM-IV (Personality Change); HADS | n=11 (33.3%) experienced personality changes |
| 65 | Nuhu & Yusuf | 2012 | Retrospective observational | n=75, m=68, age=32.2, PTA=5.45 days | Nigeria | RTA=69.3%; Other=30.7% | Premorbid personality n=71, maladjusted=5 | Not specified | Not specified |
| 66 | Rodriguez-Bailon et al. | 2012 | Cross-sectional | TBI=9, M=7, age=29.4, edu=13yrs; C=9, M=6, age=30.89, edu=14 yrs | Spain | The inclusion criterion for the frontal group was the presence of damage in the prefrontal lobe confirmed by neuroimaging and the neuropsychological assessment performed. | DSM-IV Criteria | MCMI; DSM | All the patients had impairment in the dorsolateral prefrontal cortex, the orbital prefrontal cortex or the anterior cingulate cortex. |
| 67 | Huang et al. | 2014 | Cross-sectional | B-TBI n=84, PostInj=8.7m, m=84, age=28.3.; NonB-TBI n=48; m=34; age=30.2.; Control n=79; civ=68; m=27; age=28.4 | USA | War: Blast injury (B-TBI); RTA, Sport, Falls (nonB-TBI) | HISC | GCS; HISC (Personality Change) | Not specified |
| 68 | Mathiesen et al. | 2014 | Case-control study | BPD=20, m=3, 18-50; OPD=24, m=15, 20-60 | Denmark | Not specified | OPD | DSM-IV; KAPP | Not specified |
| 69 | Pachalska et al | 2014 | Case study | Male, 27yo | Poland | Work-related accident | Post-injury: aggressiveness, anxiety, impulsiveness, inappropriateness and lack of good manners, unreasonable behaviour, hypersensitivity, and irritability. | FBInv | Frontal Lobe |
| 70 | Snell et al. | 2014 | Retrospective observational | n=14, m=12, age=41 years; follow-up= 5.8 yrs post-inj. | Australia | RTA= 9; sport-related injury = 2; fall= 2; firecracker explosion=1 | EBIQ | EBIQ | Complex fracture; 3 of the 4 axial facial levels (upper and lower maxillae, naso-orbito-zygomatic, and cranial vault) were impacted |
| 71 | Diaz et al. | 2014.1 | Case control | Total=41, m=35, age=31; PChange=14, m=13, age=33.9; nPChange=29, m= 22, age=29.8 | Brazil | nonmissile traumatic brain injury | DSM-IV | DSM-IV | Not specified |
| 72 | Diaz et al. | 2014.2 | Cohort study | N=43, m=36, age=31.6, | Brazil | Two pts trauma were not mentioned. 69.7% =RTA, 7%= assault,16.3%= falls, 2.3%= a bicycle accident. | SCID | GCS; DSM-IV; SCAT | Not specified |
| 73 | Bodley-Scott & Riley | 2015 | Qualitative | Small group n= 5; 29-42yo; 9months-7years post injury | UK | Sporting accident n=1; fall n=3; assault n=1 | "Personality changes" not defined | Interview Schedule | Not specified |
| 74 | Lauterbach et al. | 2015 | Retrospective observational | n=60; m=47, age=46, | USA | RTA; Falls | Not specified | GCS | Not specified |
| 75 | Manjila et al. | 2015 | Case study | Male, 30yo, | USA | RTA | Post-injury: emotional instability, aggressiveness, possessiveness, impulsivity. Rechristened himself. Grandiose thoughts, obsessive-compulsive traits. Took risky decisions. | Not specified | Frontal Lobe |
| 76 | Menger et al. | 2015 | Case study | Male, 41yo (inj) | USA | Work-related accident: Assassination Attempt | Not specified | Not specified | Left frontal lobe, corpus callosum, right frontal and temporal lobes. There was also a bullet fragment with an associated intraparenchymal haemorrhage in right temporal lobe. |
| 77 | Miskey et al. | 2015 | Cross-sectional | n=144, m=89.6%, age=35.29, edu=13.92. C=40; mTBI=31; H=25; mTBI/PTSD=23; mTBI/PTSD=25. | USA | Not specified | PAI | PAI | Not specified |
| 78 | Norup & Mortensen | 2015 | Cohort study | TBI=22, TBI-SO=22; C=22, C-SO=22. | Denmark | Not specified | NEO-FFI; EPQ | NEO-FFI; HRQOL; EPQ | Not specified |
| 80 | Ikram et al. | 2016 | Case study | Henry VIII |  | Multiple Sport-Related Incidents | Impulse control, irritability, anger outbursts, depression, sleeplessness, forgetfulness. | Not specified | Not specified |
| 81 | Weddell & Wood | 2016 | Cross-sectional | NHS = 40; m=30; Age=39.8; MSinceInj=64.5; ML = 31; M=22; age=33.8; MSinceInj=49.3 | UK | NHS: RTA=45%, Fall=30%, Assault=17.5%, other = 7.5%; ML: RTA=87.1%, Fall=9.6%, Assault=3.2%, other = 0.0% | Self-reported personality changes | GCS; DEX; BIS-11; WAIS; WMS; BADS; BSIT; SIT; HADS; BDI-FS; STAXI-II; Family questionnaire | Not specified |
| 82 | Wong et al. | 2016 | Cross-sectional | TBI=81, m=61, age=44.6, edu=12, AA=60 W=17 H=1; SO= 76, m=23, age=49.9, edu=12.7, AA=57 W=18 | USA | medically documented complicated-mild, moderate, or severe TBI | | BIS/BAS; PNAS; AQ; GCS; WTAR | Not specified |
| 79 | Achinivu | 2017 | Case study | Female, 58yo, inj at 54 | UK | RTA | OPD | GCS | bilateral small acute subdural hematomas and multiple contusions especially in both temporal lobes |
| 83 | Cohen-Zimerman et al. | 2017 | Cross-sectional | HC = 62.23yo; education = 15.06; N= 39. PC = 63.70yo; education = 15.45; N=33. R dlPFC = 63.27yo; education = 13.64; N=11. L dlPFC = 62.31yo; education = 13.56; N=15 | USA | Not specified | Machiavellianism | AFQT; WAIS; Mississippi PTSD scale; BDI; STAI; BNT; Faux Pas story/Control; BEES; MSCEIT; D-KEFS | Left dorsolateral prefrontal cortex |
| 84 | Margetic & Margetic | 2018 | Case study | Male, 52yo | Croatia | Work-related accident | OPD | GCS; ICD-10 (personality); WBI; WMS | a fracture of the skull base and the right temporal bone, along with temporal epidural hemorrhage |
| 85 | Weddell & Wood | 2018 | Cross-sectional | N=71, M=52, age=37.2, mntssinceinj=57.9 | UK | RTA = 60.6%, Fall = 14.1%, assault=7%, other=18.3% | Self-reported personality changes | GCS; BSIT; FSBQ; HADS, BDI-FS, STAXI-II; Family Questionnaire; WAIS, WMS; DKEFS; DEX | Not specified |
| 86 | Aldossary et al. | 2019 | Cohort study | Pts with (n=70[m=52]; 39.4 +- 9.2) vs without diffuse axonal injury (n=181[m=141]; 40.3 +- 8.4) | Saudi Arabia | Traffic (w/oDAI n=52; w/DAI n=134); Fall (w/oDAI n=10; w/DAI n=33); Pupillary Abnormality (w/oDAI n=8; w/DAI n=23); Other (w/oDAI n=8; w/DAI n=14) | "Personality changes" not defined | GOSE; SCAN (perhaps used for personality?); MMSE | Not specified |
| 87 | Merz et al. | 2019 | Cross-sectional | Total=612, m=201, age = 35.63, w=74.8%; TBI =110, time since inj=427.3 weeks | USA | Not specified | NEO-FFI | IPIP-NEO-FFI; PCSS | Not specified |
| 88 | Durcan et al. | 2020 | Case study | 46yo male | Ireland | Work-related accident | Pre-injury: affable and gregarious. Post-injury: paranoid, irritable, short-tempered; difficulties socialising, withdrew from society. Poor concentration, memory, and lost his job. Readily insults people, showing no remorse or empathy | Not specified | left fronto-parietal and temporal lobe damage. |
| 89 | McIntyre et al. | 2020 | Cross-sectional | N=89; M=39, Age=46.6 | Canada | Not specified | BFI | PHQ-9; AAQ; ASI; ADHS; BFI; BCOPE; RSES; GCS | Not specified |
| 90 | Balan et al. | 2021 | Cross-sectional | Total n=41 [m=34]; 32 avg. No R2W = n=21 [m=18]; 35 avg. R2W = 20 [m=16]; 29.5 avg. | Brazil | TBI | "Personality changes" not defined | DSM-5 ("personality change"); Overt Aggression Scale; Neuropsychiatric Inventory-Questionnaire; Starkstein Apathy Scale | Not specified |
| 91 | Banerjee et al. | 2021 | Case study | 25yo female (RTA age 23) | India | RTA | Post-injury changes included: Increased irritability, anger outbursts, stubbornness and rigidity, an inability to experience her sense of ‘self’, uncomfortable in social situations, inability to engage in conversations, or social interactions, unable to comprehend humour, sarcasm, pun, metaphors. Premorbid Personality; fun-loving extrovert, good at extra-curricular activities. K.S. found it difficult to relate to this change. | NIMHANS neuropsychological battery; Beck’s Depression Inventory | K.S often reported how ‘she is not the same following the injury’. |
| 92 | Li et al. | 2021 | Cross-sectional | TBI=234, m=166, age=44.43; C=277, male = 183, age = 46.03 | China | Not specified | ICD-10 criterion F07 and Z87.820 | ADL scale, SDSS scale, and scale of personality change following a traumatic brain injury | Not specified |
| 93 | Mundluru et al. | 2021 | Cross-sectional | Total = 1774, m=612 (age=68.7 edu=16.4), f=1162 (age=69.8 edu=16.3); TBI=266 (M=118, age=68.7, edu=16.4); C=1508 (m=494, age=69.8, edu=16.3) | Canada | Not specified | NPI-Q | NPI-Q | Not specified |
| 94 | Brett et al. | 2022 | Cross-sectional | Total = 106; m=73, age=21.37, ethnicity B=8, W=97, O=1 | USA | Sport-Related Incident | Big Five Inventory | BFI; BSI-GSI; MPQ; PSQI; SCAT; WTAR | Not specified |
| 96 | Guzowski et al. | 2022 | Cohort study | Athlete n=100: Male n=100, Age=18, Race white=75, black=23, other=2; CTBI n=75: Male=35, age=49.7, Race white=50, black=18, other=7; Control n=79: Male=56, age=48, Race white=57, black=18, other=5 | USA | Sport-Related Incident | positive and negative emotionality; negative affect, detachment, antagonism, disinhibition, and psychoticism | MPQ; DSM-5; DIS-11 | Not specified |
| 97 | Li et al. | 2022 | Cross-sectional | OPD=340, m=198, age=41.4, edu=8.8; nOPD=687, m=363, age=37.9, edu=11.3 | China |  | Neuroticism Extraversion Openness Five-Factor Inventory | NEO-FFI; ICD-10 | Not specified |
| 39 | Belfry et al. | 2023 | Cross-sectional | n=637, m=100%, age=m35 | Canada | ABI based on several coded variables regarding events consistent with the potential to cause brain injury documented in the medical file | Personality disorder TBI=36 (28%) C=73 (14%); Antisocial personality disorder TBI= 22 (17%), C=51 (10%) | non-standardised; VRAG; CATS | Not specified |
| 98 | Ling et al. | 2023 | Cross-sectional | TBI n=31, Controls n=24; TBI: M age=36.90 (SD=10.81), 14 male/17 female, edu=12.81y (SD=2.50); Controls: M age=32.96 (SD=12.10), 15 male/9 female, edu=14.77y (SD=3.66) | China | Neurocognitive disorder after TBI diagnosed per ICD-10; Patients recovered for at least 6 months after TBI; Received clinical care | Scale of personality change following TBI | EEG contingent negative variation correlated with personality change | Not specified |
| 95 | Filipčíková et al. | 2024 | Cross-sectional | 25 TBI (19M, 6F); 25 controls (18M, 7F). TBI group: mean age 46.76 (SD=15.78), mean education 12.76 years (SD=2.31), average time since injury 14.67 years (SD=11.67) | Australia | Severe TBI: PTA > 7 days, or ≥1 day altered consciousness, or GCS 3-8. Median PTA: 52 days (IQR=53, range 9-180). Average age at injury: 31.54 years (SD=15.55, range 13-59). At least 1 year post-injury. | Social disinhibition and aggression components (physical aggression, verbal aggression, anger, hostility). Higher self-reported anger associated with higher self-reported social disinhibition, controlling for age and mood. | Frontal Systems Behaviour Scale (FrSBe) - Disinhibition subscale; Social Disinhibition Interview (SDI); Buss-Perry Aggression Questionnaire (BPAQ) - Physical, Verbal, Anger, Hostility subscales | Not specified |
| 99 | Edwards et al. | 2024 | Cross-sectional | 328 US service members and veterans: Uncomplicated mild TBI (MTBI, n=155), Complicated mild/moderate/severe TBI (STBI, n=97), Non-injured controls (NIC, n=76). Three cohorts by time since injury: ≤12 months (MTBI n=46, STBI n=41), 3-5 years (MTBI n=57, STBI n=30), 8-10 years (MTBI n=52, STBI n=26). Predominantly male (87-100% in TBI groups). Mean age at assessment ranged 29.5-40.3 years across cohorts. | USA | TBI severity classified: (1) Uncomplicated mild: GCS 13-15, PTA <24h, LOC <30min, no intracranial abnormality; (2) Complicated mild: same criteria but with intracranial abnormality on CT/MRI; (3) Moderate: LOC >30min-24h, PTA 1-7 days; (4) Severe: LOC >24h, PTA >7 days. Combat or non-combat related. Part of DVBIC-TBICoE 15-Year Longitudinal TBI Study. Cross-sectional assessment at three time points post-injury. | Higher MMPI-2-RF scores associated with lower biomarker concentrations (GFAP, NfL, tau) in subacute/chronic phase up to 5 years post-injury, with reverse trend at 8-10 years. proneness (STBI ≤12mo), head pain (STBI 8-10yr). Lower NfL related to demoralization, somatic/neurological/cognitive complaints (MTBI ≤12mo), demoralization (STBI ≤12mo, 3-5yr), head pain, stress/worry (STBI 3-5yr). At 8-10 years, increased tau associated with worse demoralization and anxiety (STBI). Findings suggest biomarkers may relate to neurobehavioral outcomes throughout recovery trajectory. | Minnesota Multiphasic Personality Inventory-2-Restructured Format (MMPI-2-RF) - 11 scales: | Not specified |
| 100 | Hanna et al. | 2024 | Case study | 1 case: 63-year-old male with history of hypertension, chronic kidney disease on dialysis, and TBI. Injured at age 18. No family history of schizophrenia or psychosis. Lives alone, able to perform activities of daily living. Adherent to medications. | USA | Severe TBI from gunshot wound through right frontal bone at age 18. Pre-injury: patient was very social, smart, had many friends. Injury resulted in cognitive dysfunction and seizures. History of inpatient psychiatric admissions | Personality changes included: impulsivity, excessive spending (gambling, buying unnecessary things), development of short temper. At presentation: depressed mood, aggressive behaviors, suicidal ideation, internally preoccupied with disorganized thoughts. Developed schizophrenia over several years following trauma. | Not specified | Encephalomalacia in right frontal convexity region. Deformity in calvarium just above right frontal convexity region. |
| 101 | Riccitelli et al. | 2024 | Case study | 1 case: 34-year-old right-handed woman. Sustained severe head trauma at age 18. 16 years post-injury | Switzerland | Severe brain injury from road traffic accident at age 18. Cranial trauma induced coma due to bilateral subdural hematomas requiring emergency craniotomy. Widespread axonal damage in subcortical white matter. | Complex psychopathology including: impulsivity leading to destructive behavior and self-harm, delusions, mood instability with depressive episodes, emotional suppression, obsessive-compulsive behaviors. Severity often required emergency psychiatric interventions and compulsory hospital admissions. Treatment: 20 daily sessions of dual-site sequential rTMS (inhibitory 1Hz right DLPFC, excitatory 10Hz left DLPFC). Results - 2 weeks: decreased impulsivity (6.4%), significant reduction in OCD symptoms (52.4%, compulsions 91.7%), improvements in attention and processing speed (18.7%), improved inhibitory control (34.6%). 4 weeks: further impulsivity decline (17.7%), attentive impulsiveness improved (26.3%). 8 weeks follow-up: persistent positive effects including enhanced positive emotions. Daily emotions diary showed qualitative increase in positive emotions (baseline 3.9 to 6.5 at 8 weeks) and decrease in negative emotions (baseline 3.9 to 1.9 at 8 weeks). CGI severity improved from severe (6) at baseline to moderate (4) at 4 weeks. | Barratt Impulsiveness Scale (BIS-11) for impulsivity with subscales (attentional, motor, non-planning). | Bilateral subdural hematomas (requiring craniotomy). Widespread axonal damage at subcortical level primarily in: frontotemporal regions (predominantly left), temporobasal regions (predominantly left), trunk of corpus callosum, right upper paravermial region. MRI showed hemorrhagic contusions at: left forceps minor, left dorsolateral prefrontal cortex, right inferior frontal gyrus, splenium of corpus callosum, periaqueductal mesencephalon tegmental region. Follow-up MRI showed severe lateral ventricular and aqueductal enlargement related to parenchymal volume loss. |

**Table S3.** Meta-regression results

| **Moderator** | **k** | **Estimate** | | **CI** | | **p** |
| --- | --- | --- | --- | --- | --- | --- |
| *Broad personality change* | | | | | | |
| Mean age | 8 | -0.004 | | [-0.025, 0.017] | | 0.724 |
| % Female | 8 | 0.002 | | [-0.007, 0.01] | | 0.714 |
| Follow-up (months) | 8 | 0.003 | | [0, 0.005] | | 0.044* |
| % mild TBI | 6 | NA | | [NA, NA] | | NA |
| % moderate TBI | 6 | 0.007 | | [-0.002, 0.017] | | 0.104 |
| % severe TBI | 6 | -0.007 | | [-0.017, 0.002] | | 0.104 |
| Year of publication | 9 | -0.006 | | [-0.016, 0.003] | | 0.186 |
| Risk of bias (%) | 9 | -0.004 | | [-0.009, 0.001] | | 0.116 |
| *Secondary personality disorder diagnosis* | | | | | | |
| Mean age | 8 | -0.004 | | [-0.011, 0.003] | | 0.293 |
| % Female | 8 | -0.001 | | [-0.005, 0.002] | | 0.441 |
| Follow-up (months) | 7 | 0 | | [-0.001, 0] | | 0.205 |
| % mild TBI | 8 | -0.002 | | [-0.005, 0.001] | | 0.196 |
| % moderate TBI | 8 | -0.007 | | [-0.013, -0.001] | | 0.018* |
| % severe TBI | 8 | 0.002 | | [0, 0.004] | | 0.093 |
| Year of publication | 8 | 0.007 | | [0.004, 0.011] | | <0.0001* |
| Risk of bias (%) | 8 | 0 | | [-0.003, 0.003] | | 0.813 |
| *Entire sample* | | |  | |  | |
| Mean age | 16 | -0.008 | | [-0.022, 0.005] | | 0.219 |
| % Female | 16 | 0 | | [-0.006, 0.006] | | 0.976 |
| Follow-up (months) | 15 | 0 | | [-0.002, 0.001] | | 0.79 |
| % mild TBI | 14 | -0.007 | | [-0.013, -0.001] | | 0.023* |
| % moderate TBI | 14 | 0.003 | | [-0.005, 0.012] | | 0.445 |
| % severe TBI | 14 | 0.002 | | [-0.003, 0.007] | | 0.378 |
| Year of publication | 17 | -0.008 | | [-0.017, 0.001] | | 0.093 |
| Risk of bias (%) | 17 | -0.005 | | [-0.009, 0] | | 0.037* |
